# The Associations of Mental Disorders in Children With Parents’ Subsequent Mental Disorders: A Nationwide Cohort Study From Finland and Denmark

**DOI:** 10.1101/2024.05.31.24308106

**Authors:** Christian Hakulinen, Mai Gutvilig, Ripsa Niemi, Natalie C. Momen, Laura Pulkki-Råback, Petri Böckerman, Oleguer Plana-Ripoll, Kaisla Komulainen, Marko Elovainio

## Abstract

**Background:** Intergenerational transmission of mental disorders has been well established, but it is unclear whether exposure to a child’s mental disorder increases parents’ subsequent risk of mental disorders.

**Aims:** We examined the association of mental disorders in children with their parents’ subsequent mental disorders.

**Methods:** In this population based-cohort study, we included all persons with children born in Finland or Denmark in 1990–2010. Information of mental disorder was acquired from national registers. The follow-up period began when the parent’s eldest child was 5 years old (for codes F10–F60 ICD-10) or 1 year old (for codes F70–F98 of the ICD-10) and ended on December 31^st^, 2019, or when the parent received a mental disorder diagnosis, died, or emigrated from Finland or Denmark. The association of mental disorders in children with their parents’ subsequent mental disorders was examined using Cox proportional hazards models.

**Results:** The study cohort included 1 651 723 parents. In total, 248 328 women and 250 763 men had at least one child who was diagnosed with a mental disorder. The risk of a parent receiving a mental disorder diagnosis was higher among those who had a child with a mental disorder compared to those who did not. For both parents, the hazard ratios were greatest in the first six months after the child’s diagnosis (HR between 2.04-2.54), followed by a subtle decline in the risk (after two years, HR between 1.33-1.77).

**Conclusion:** Mental disorders in children were associated with a greater risk of subsequent mental disorders among their parents. Additional support is needed for parents whose children have been recently diagnosed with a mental disorder.

## Introduction

It is well recognized that mental disorders cause a substantial burden on family members, and especially on parents.^1,2^ This mental and emotional toll may stem from many different sources. Mental disorders have considerable heritability.^3,4^ and, due to intergenerational mechanisms operating through genetic and environmental factors, both parents and their children might suffer from poor mental health.^5,6^ On the other hand, parents exposed to a child’s mental disorder may experience distress arising from witnessing their own child’s disorder, which is often accompanied by a need to invest additional economic resources and social support to provide care for their child. Each of these can adversely affect the well-being of the caregiver and other family members.^7–15^

The association between children’s mental disorders and parents’ mental health has been traditionally investigated from the perspective of caregivers, i.e., persons taking care of a family member with a mental disorder. Most previous studies are based on relatively small and nationally non-representative samples,^10–12,15,16^ focusing specifically on those individuals who are in practice taking care of the persons with a mental disorder, and thus, these studies do not describe the overall impact of mental disorders on both parents. Although some studies have explicitly examined the consequences of children’s mental disorders for their parents, they have focused on specific disorders, such as schizophrenia^17^ and autism^18^ in children, and large-scale nationwide investigations across the full spectrum of mental disorders are lacking. Using nationwide register data from Finland and Denmark, we examined whether mental disorders in children are associated with their parents’ subsequent risk of mental disorders. Whereas previous studies focused only on few disorders and specifically on caregivers, we examined the population-level associations across the full spectrum of mental disorders diagnosed in childhood, adolescence, and early adulthood. Interactive visualizations of the results are available at https://mentalnet.shinyapps.io/app_morbidity/.

## Methods

### Study Population

The study population consisted of all individuals whose children were born in Finland or Denmark between 1990 and 2010. Individuals with children who had died or emigrated from their country of birth before their first birthday were excluded. The datasets were constructed by compiling nationwide registries (Supplement for details), which were linked using unique personal identification numbers.

The Ethics Committee of the Finnish Institute for Health and Welfare approved the study plan (THL/184/6.02.01/2023§933). Data were linked with the permission of Statistics Finland (TK-53-1696-16), Statistics Denmark, and the Finnish Institute of Health and Welfare. According to Finnish and Danish law, informed consent is not required for register-based studies. The present study adhered to the Strengthening the Reporting of Observational Studies in Epidemiology (STROBE) reporting guideline.

### Study Design

Parental risk of receiving a mental disorder diagnosis relative to having a child with a mental disorder diagnosis was examined. The follow-up period began when the parent’s eldest child was deemed sufficiently old to be at risk of receiving a mental disorder diagnosis (5 years old for codes F10–F60 of the International Classification of Diseases, 10th revision (ICD-10); 1 year old for codes F70–F98 of the ICD-10). Follow-up ended on December 31^st^, 2019, or when the parent received a mental disorder diagnosis, died, or emigrated from Finland or Denmark. As the focus of the study was on new-onset mental disorders, parents with a history of diagnosed mental disorders prior between January 1, 1970 and the start of follow-up were excluded.

### Measurement of Mental Disorders

In Finland, mental disorders were classified using the International Classification of Diseases, 8th revision (ICD-8) from 1970 to 1986, ICD-9 from 1987 to 1995, and ICD-10 from 1996 onwards. The International Classification of Primary Care, 2nd edition (ICPC-2) was used alongside the ICD-10 in primary healthcare data and the ICPC-2 mental health-related diagnoses were converted to corresponding ICD-10 sub-chapter categories.^19^ In Denmark, ICD-8 was used from 1969 to 1993 and ICD-10 from 1994 onwards.

Exposure to a child’s mental disorder was defined using ICD-10 codes (and corresponding ICD-8/ICD-9/ICPC-2 codes) and classified into nine broad categories of mental disorders based on subchapter F-categories of the ICD-10 (e.g., F10–F19: Mental and Behavioral Disorders Due to Psychoactive Substance Use). For full details on diagnostic categories and the specific disorders included in each category (**Table S1** in Supplement).

The primary exposure was any mental disorder of a child, defined as the first recorded ICD- 10 subchapter F code (F00–F99, or corresponding ICD-8/ICD-9/ICPC-2 codes) among children. Exposure to any mental disorder of a child was restricted to diagnoses received when the child was 1–25 years old. The date of a child’s diagnosis was considered the date of a parent’s exposure. In addition, exposure to specific mental disorders of a child was examined, which was restricted to diagnoses received when the child was 5–25 years old (F10–F60) or 1–25 years old (F70–F99). The date of a child’s first diagnosis in each diagnostic category was considered the date of a parent’s exposure to that disorder. One child could act as the exposure for several mental disorders if they had diagnoses belonging to more than one diagnostic category. In the case of several children of the same parent receiving a mental disorder diagnosis within the same disorder category, the date of the earliest diagnosis among all their children was considered the date of exposure to that disorder.

The outcome of the study was time to a parental mental disorder. The emergence of a parental mental disorder was defined as the first recorded ICD-10 subchapter F code (or its corresponding ICD-8/ICD-9/ICPC-2 code) after the start of follow-up.

### Covariates

Parent’s education was measured at the beginning of follow-up and further aggregated into the following three categories: primary education, secondary education, and higher education. Parent’s age at the birth of their first child was categorized into the subsequent seven categories: below 20, 20–24, 25–29, 30–34, 35–39, 40–44, and above 45 years.

### Statistical analyses

The hazard ratios (HR) of a parent’s mental disorder diagnosis in relation to the parent having a child with a mental disorder were examined using Cox proportional hazards models with time since the beginning of follow-up as the underlying time scale. The follow-up period began when the parent’s eldest child was either 5 years old (when ICD-10 diagnoses F10–F60 were used as an exposure) or 1 year old (when ICD-10 diagnoses F70–F98 were used as an exposure). Exposure to a child’s mental disorder was treated as time-varying, resulting in each parent being considered unexposed until their child received a mental disorder diagnosis. Moreover, to account for the HR of a parent’s mental disorder depending on time since the child’s mental disorder, time-dependent HRs were modelled with five different time periods after the child’s diagnosis examined (>0–6 months, >6–12 months, >1–1.5 years, >1.5–2 years, or >2 years). When hazards were not proportional, we interpreted the HRs as average ratios over the time period.^20^ Separate models were estimated for men and women and different categories of a child’s mental disorder. Finnish and Danish data were analyzed separately using the same analytical strategy.

All models were adjusted for the year the follow-up began to account for calendar time. Additionally, models were adjusted for the parent’s age at the birth of their first child and the parent’s educational attainment at the beginning of follow-up. When examining exposure to specific mental disorder categories, analyses with additional adjustment for prior exposure to each mental disorder category except the exposure at hand were also conducted. For example, when examining exposure to a child’s mood disorder, a child of the same parent could have received an anxiety diagnosis prior to the date of the mood disorder. This was adjusted for in the comorbidity adjusted models. For further details on comorbidity adjustment, see Supplement.

As a sensitivity analysis, we gauged patterns in parents’ HRs prior to the child’s diagnosis to examine whether temporal patterns could be confounded by natural changes due to aging or shared genetic and environmental factors rather than reflecting associations with the onset of the child’s mental disorder. In addition to the aforementioned five time periods after the child’s diagnosis, two new time periods were examined: 12–6 months prior to the child’s diagnosis and the 6 months preceding the child’s diagnosis. Importantly, the study population in the sensitivity analyses differed slightly from the main models as the beginning of follow-up was moved back by a year, resulting in partly different people being excluded from the study population. Last, we replicated the main analyses using only Finnish secondary care data.

All data were analyzed using Stata version 17.0 (StataCorp) and R version 4.2.2 (R Core Team, 2022) employing packages gtsummary (v. 1.6.2), tidyverse (v.1.3.2), and shiny (v.1.7.5).^21–23^

## Results

The number of men and women at risk of receiving a new-onset mental disorder at the start of follow-up and those excluded due to a mental disorder diagnosis preceding the start of follow-up are reported in **Table 1**. Overall, 248 328 women and 250 763 men had at least one child who was diagnosed with a mental disorder. The baseline characteristics of the study population in Finland and Denmark are shown in **Table 2**.

**Table 1.**
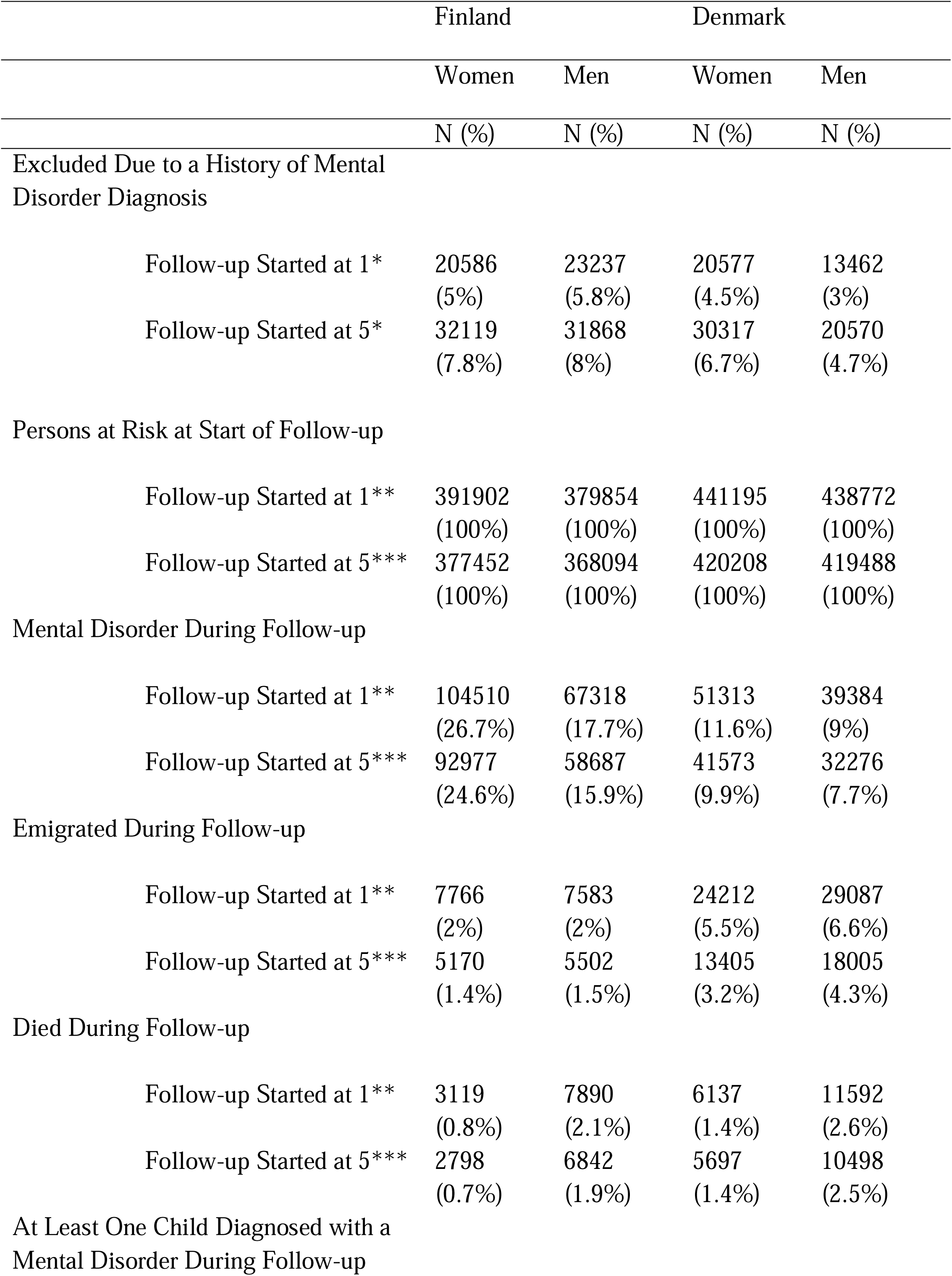
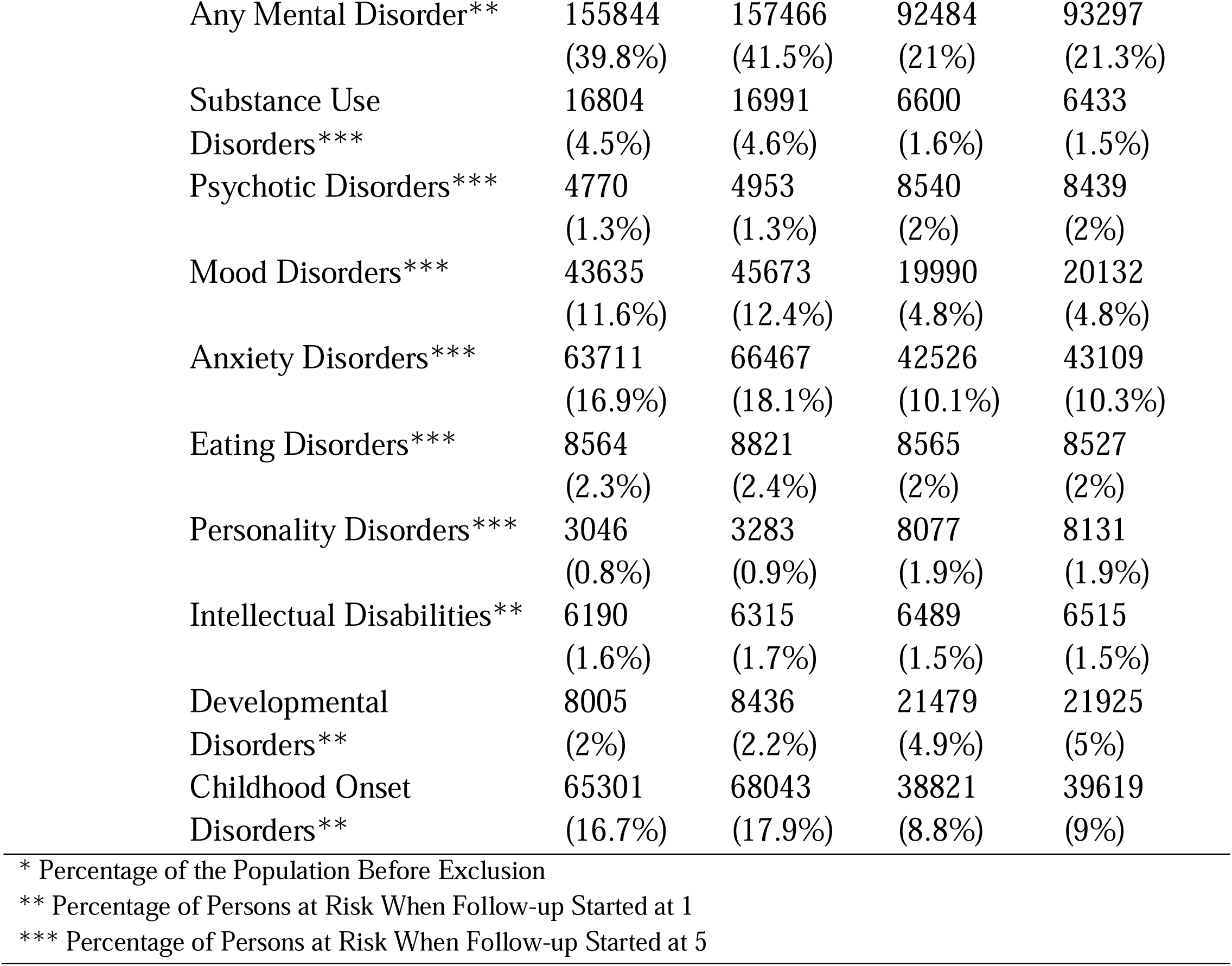
Frequencies of Persons Excluded Due to a History of Mental Disorder Diagnosis, at Risk at Start of Follow-up, Diagnosed or Otherwise Censored During Follow-up, and Exposed to at Least One Child Diagnosed with a Mental Disorder During Follow-up.

|  | Finland |  | Denmark |  |
| --- | --- | --- | --- | --- |
|  | Women | Men | Women | Men |
|  | N (%) | N (%) | N (%) | N (%) |
| Excluded Due to a History of Mental Disorder Diagnosis |  |  |  |  |
| Follow-up Started at 1* | 20586<br>(5%) | 23237<br>(5.8%) | 20577<br>(4.5%) | 13462<br>(3%) |
| Follow-up Started at 5* | 32119<br>(7.8%) | 31868<br>(8%) | 30317<br>(6.7%) | 20570<br>(4.7%) |
| Persons at Risk at Start of Follow-up |  |  |  |  |
| Follow-up Started at 1** | 391902<br>(100%) | 379854<br>(100%) | 441195<br>(100%) | 438772<br>(100%) |
| Follow-up Started at 5*** | 377452<br>(100%) | 368094<br>(100%) | 420208<br>(100%) | 419488<br>(100%) |
| Mental Disorder During Follow-up |  |  |  |  |
| Follow-up Started at 1** | 104510<br>(26.7%) | 67318<br>(17.7%) | 51313<br>(11.6%) | 39384<br>(9%) |
| Follow-up Started at 5*** | 92977<br>(24.6%) | 58687<br>(15.9%) | 41573<br>(9.9%) | 32276<br>(7.7%) |
| Emigrated During Follow-up |  |  |  |  |
| Follow-up Started at 1** | 7766<br>(2%) | 7583<br>(2%) | 24212<br>(5.5%) | 29087<br>(6.6%) |
| Follow-up Started at 5*** | 5170<br>(1.4%) | 5502<br>(1.5%) | 13405<br>(3.2%) | 18005<br>(4.3%) |
| Died During Follow-up |  |  |  |  |
| Follow-up Started at 1** | 3119<br>(0.8%) | 7890<br>(2.1%) | 6137<br>(1.4%) | 11592<br>(2.6%) |
| Follow-up Started at 5*** | 2798<br>(0.7%) | 6842<br>(1.9%) | 5697<br>(1.4%) | 10498<br>(2.5%) |
| At Least One Child Diagnosed with a Mental Disorder During Follow-up |  |  |  |  |
| Any Mental Disorder** | 155844<br>(39.8%) | 157466<br>(41.5%) | 92484<br>(21%) | 93297<br>(21.3%) |
| Substance Use Disorders*** | 16804<br>(4.5%) | 16991<br>(4.6%) | 6600<br>(1.6%) | 6433<br>(1.5%) |
| Psychotic Disorders*** | 4770<br>(1.3%) | 4953<br>(1.3%) | 8540<br>(2%) | 8439<br>(2%) |
| Mood Disorders*** | 43635<br>(11.6%) | 45673<br>(12.4%) | 19990<br>(4.8%) | 20132<br>(4.8%) |
| Anxiety Disorders*** | 63711<br>(16.9%) | 66467<br>(18.1%) | 42526<br>(10.1%) | 43109<br>(10.3%) |
| Eating Disorders*** | 8564<br>(2.3%) | 8821<br>(2.4%) | 8565<br>(2%) | 8527<br>(2%) |
| Personality Disorders*** | 3046<br>(0.8%) | 3283<br>(0.9%) | 8077<br>(1.9%) | 8131<br>(1.9%) |
| Intellectual Disabilities** | 6190<br>(1.6%) | 6315<br>(1.7%) | 6489<br>(1.5%) | 6515<br>(1.5%) |
| Developmental Disorders** | 8005<br>(2%) | 8436<br>(2.2%) | 21479<br>(4.9%) | 21925<br>(5%) |
| Childhood Onset Disorders** | 65301<br>(16.7%) | 68043<br>(17.9%) | 38821<br>(8.8%) | 39619<br>(9%) |

\* Percentage of the Population Before Exclusion
\*\* Percentage of Persons at Risk When Follow-up Started at 1
\*\*\* Percentage of Persons at Risk When Follow-up Started at 5

**Table 2.**
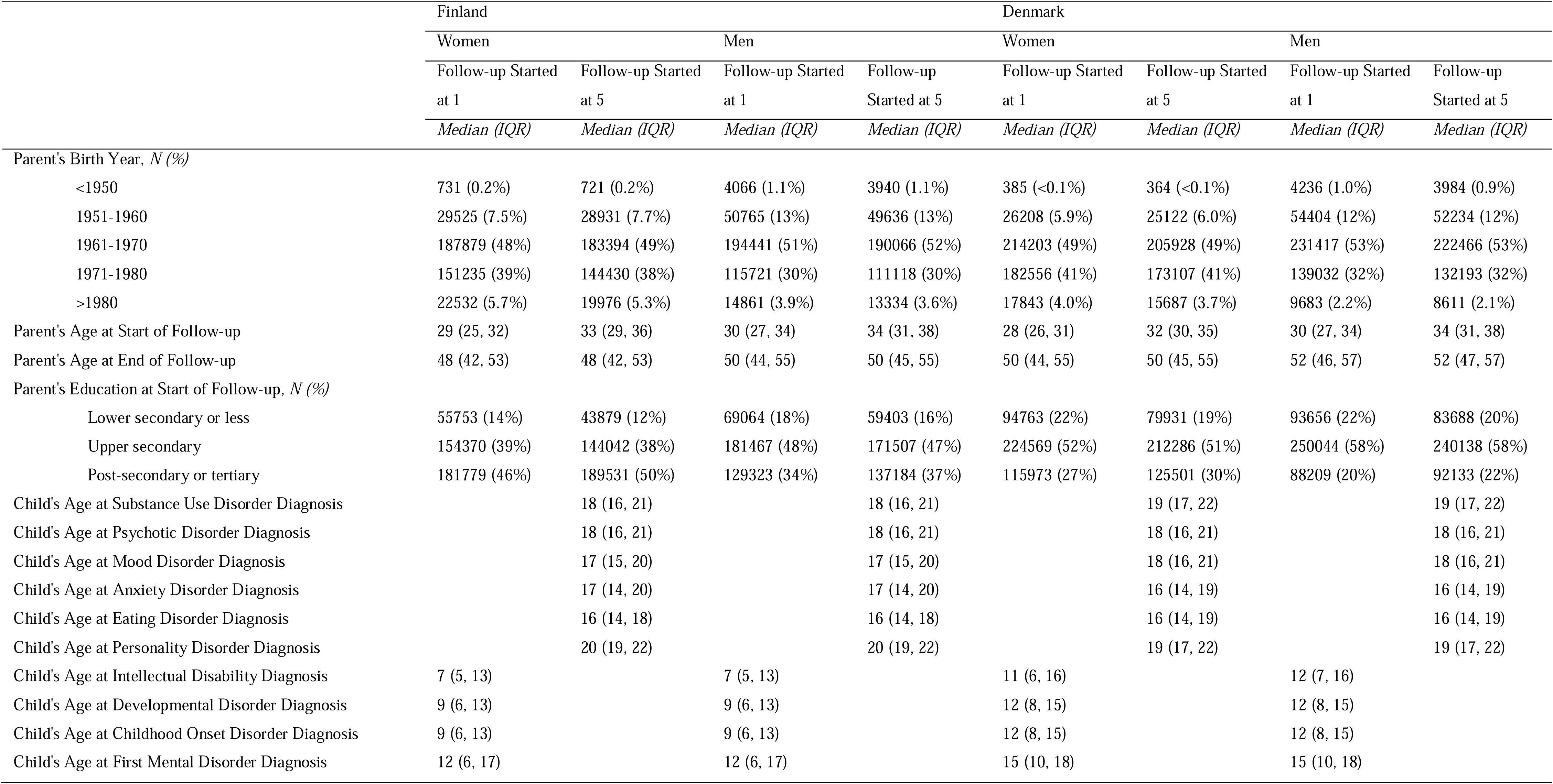
Descriptives of Study Population.

The time-dependent HRs of a parent receiving a mental disorder diagnosis depending on time since a child received a diagnosis of any mental disorder are shown in **Figure 1**. The rate of receiving a mental disorder diagnosis was higher in parents who had a child with a mental disorder compared to those who did not. Overall, the HR was higher for women than men.

**Figure 1.**
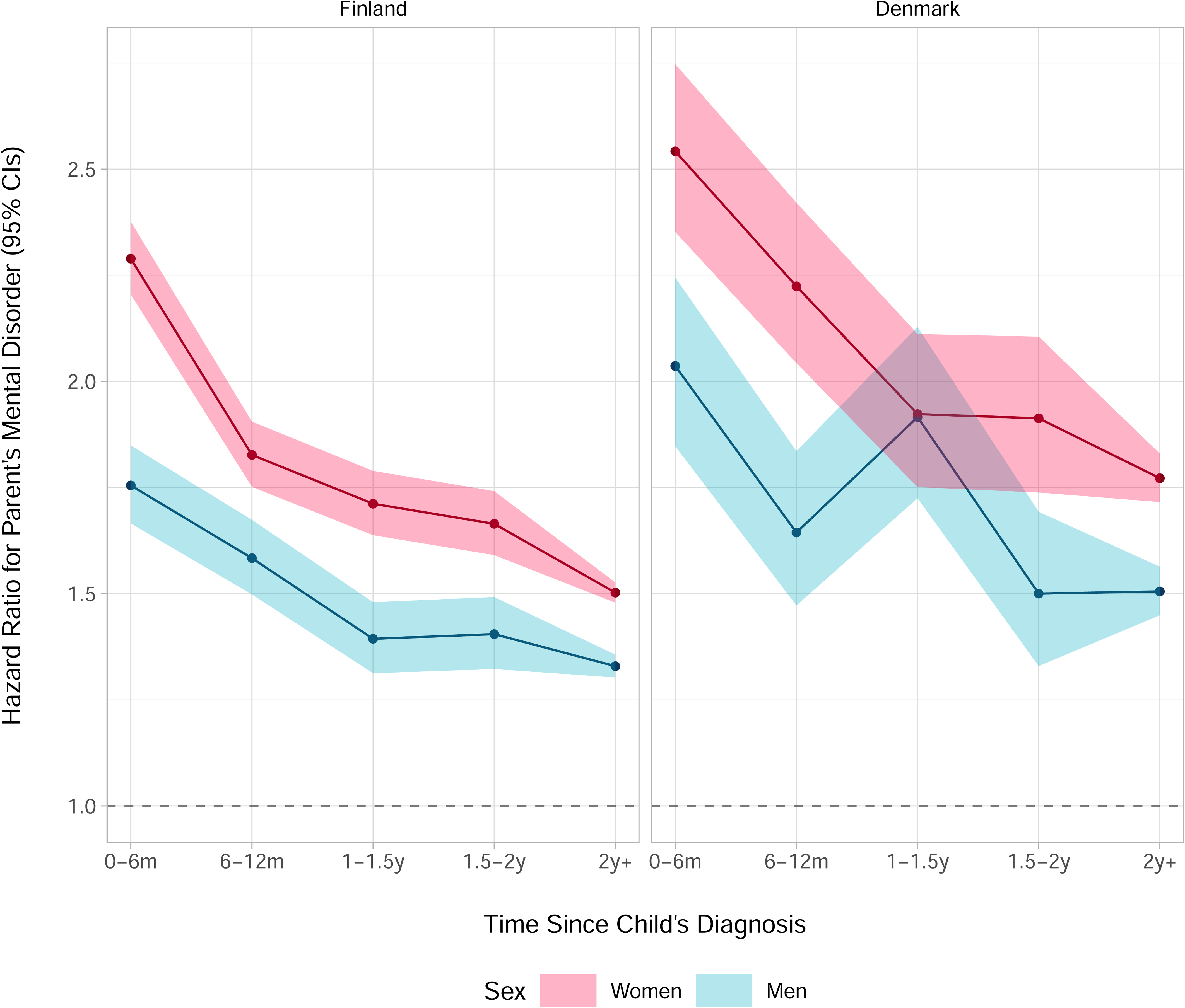
Hazard Ratios and 95% Confidence Intervals of Parents’ Mental Disorder in Relation to Any Mental Disorder of a Child According to Time Since the Child’s Diagnosis

Compared to parents without a mental disorder, the HRs for parents who had a child with a mental disorder were at their highest in the first six months following the child’s diagnosis over the follow-up. This was observed for both sexes (Finland, women: HR, 2.29 [95% CI, 2.21–2.38], men: HR, 1.75 [95% CI, 1.67–1.85]; Denmark, women: HR, 2.54 [95% CI, 2.35–2.75], men: HR, 2.04 [95% CI, 1.85–2.24]). They then gradually declined among women in both Finland and Denmark, and among men in Finland. In Denmark, the HRs among men decreased during 6–12 months following the child’s diagnosis, then increased between 1 to 1.5 years (HR, 1.92 [95% CI, 1.72–2.13]), and then again decreased. Regardless of temporal trends, the HRs remained positive and significant after two years since the child’s diagnosis across both sexes in Finland and Denmark.

Figure 2 shows the comorbidity adjusted HRs of women receiving any mental disorder diagnosis in relation to their child receiving a specific mental disorder diagnosis. Notable exceptions from the general trend – where the HRs were most elevated in the year following the child’s diagnosis followed by a subtle decline during the follow-up – were personality disorders in both Finland and Denmark, and eating and developmental disorders in Denmark, where no time-dependent associations between child’s and parent’s mental disorders were observed. Overall, the time-dependent associations were more pronounced among women in Finland than Denmark. Compared to the minimally adjusted models, the point estimates of the comorbidity adjusted models were generally lower (**Supplement Table 2**).

**Figure 2.**
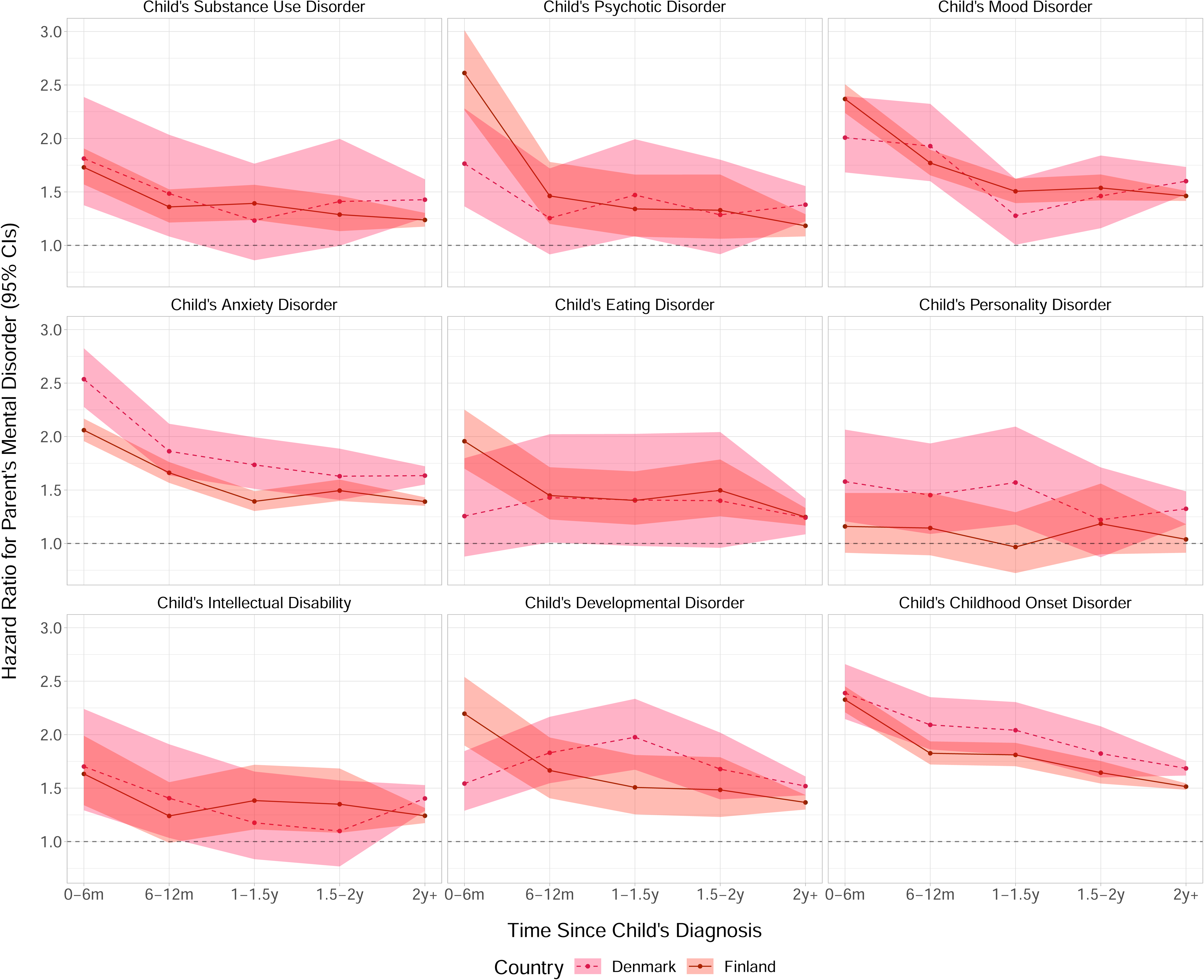
Hazard Ratios and 95% Confidence Intervals of Women’s Mental Disorder in Relation to a Child’s Specific Mental Disorder According to Time Since the Child’s Diagnosis

For men, the HRs for receiving a mental disorder diagnosis in relation to their child having a specific mental disorder are shown in Figure 3. A time-dependent association where the HRs were at their highest in the 6 months following a child’s diagnosis and then declined was consistently observed for parents of children diagnosed with anxiety disorders in both Finland and Denmark. Otherwise, the temporal patterns were inconsistent and differed between Finland and Denmark, although point estimates and confidence intervals were largely overlapping. The estimates from the comorbidity adjusted models were generally lower than the estimates from the minimally adjusted models (**Supplement Table 3**).

**Figure 3.**
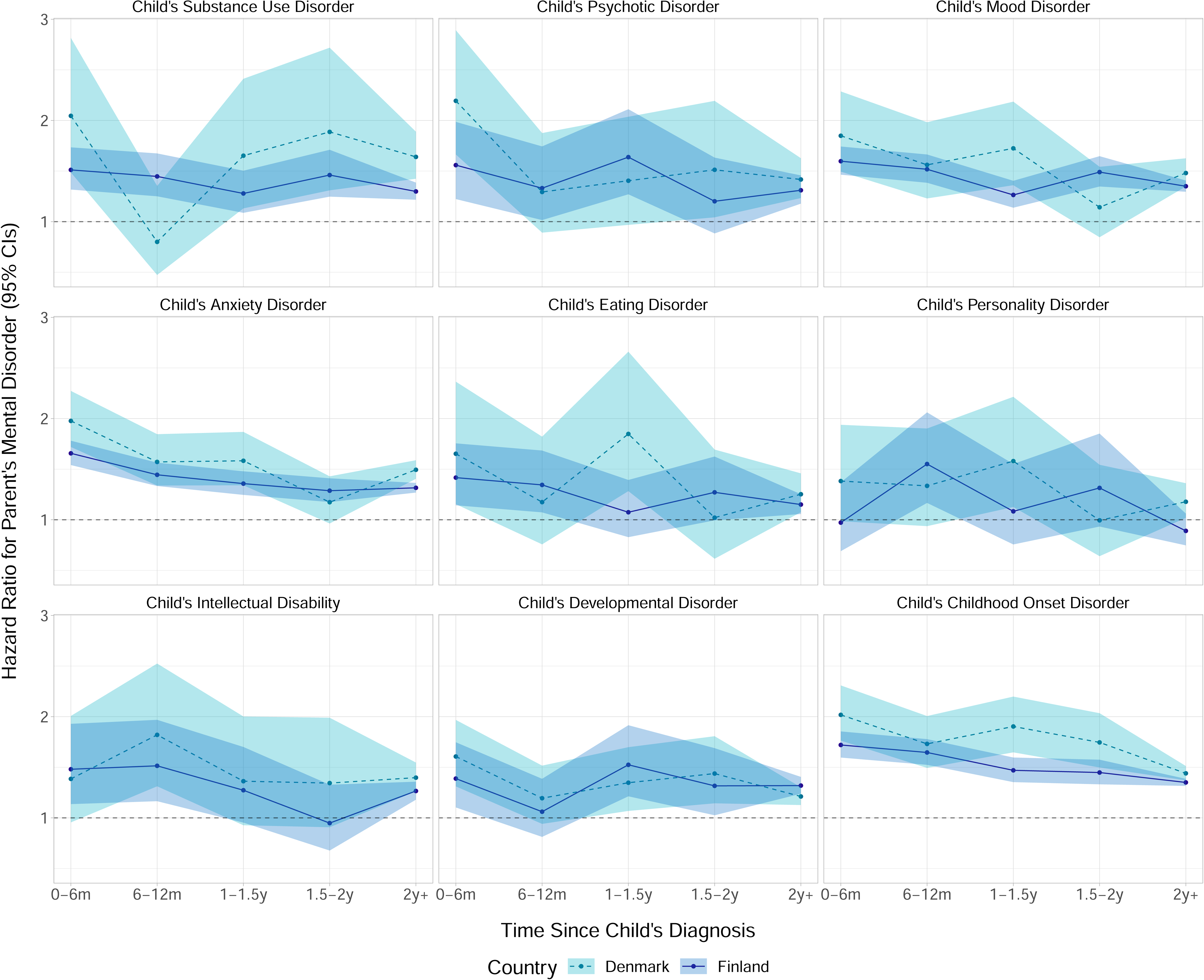
Hazard Ratios and 95% Confidence Intervals of Men’s Mental Disorder in Relation to a Child’s Specific Mental Disorder According to Time Since the Child’s Diagnosis

Details on the size and characteristics of the study population used in the sensitivity analyses are reported in **Supplement Table 4** and **Supplement Table 5**. In the sensitivity analyses, the HR of a parent receiving a mental disorder diagnosis was already higher 12 to 6 months prior to the child’s diagnosis date (**Supplement** Figure 1). The HRs increased sharply in the 6 months before the child’s diagnosis, reaching levels comparable to those observed in the 6 months post-diagnosis. The comorbidity adjusted HRs for receiving a mental disorder diagnosis in relation to a child’s specific mental disorder are shown in **Supplement** Figure 2 (**Supplement Table 6**) for women and **Supplement** Figure 3 (**Supplement Table 7**) for men. Among women, an increase in HRs in the year preceding the child’s diagnosis was observed across most diagnostic categories. Among men, the temporal patterns were largely inconsistent and varied between Finland and Denmark. Finally, in the sensitivity analyses using only Finnish secondary health care data, the HRs of a parent receiving a mental disorder diagnosis were higher than when the primary care data was also included (Supplement Figure 4). The differences were observed especially among women (**Supplement** Figure 4), and they were visible in mood, anxiety, developmental and childhood onset disorders (**Supplement** Figure 5). For men, the diagnosis specific results were more mixed (**Supplement** Figure 6).

## Discussion

In this study using two nationwide cohorts with over 1.6 million parents from Finland and Denmark, we found that the risk of a parent receiving a mental disorder diagnosis was higher among those who had a child with a mental disorder compared to those who did not. Overall, the risk was at its highest in temporal proximity to the child’s mental disorder diagnosis among both women and men, and then declined over time. When examining specific disorders of a child, a similar time-dependent trend was generally observed across most diagnostic categories in women, while the temporal patterns in men were less consistent.

To our knowledge, the present study is the most comprehensive investigation on how children’s mental disorders are associated with those of their parents. Although several studies have linked children’s mental disorders with parents’ mental health and overall well-being,^7–12^ most previous studies were based on small samples^10–12,15,16^ or focused on specific disorders.^17,18^ Utilization of nationwide registers provided comprehensive information of two entire populations, allowing us to examine the relationship between mental disorders in children and their parents with a greater precision than previously.

Having a child diagnosed with any mental disorder was consistently associated with a greater risk of a parent receiving a subsequent mental disorder diagnosis throughout the follow-up.

To a large extent, this is likely explained by genetic and environmental factors shared between children and their parents. However, we excluded parents with pre-existing mental disorders, and the markedly higher relative risk observed in temporal proximity of the child’s diagnosis, which then started to decline, suggests that the onset of a child’s mental disorder is associated with a transient increase in parents’ risk of a mental disorder. The increase was already observed six months prior to the child’s diagnosis, which is not unexpected given the diagnostic delay associated with mental disorders^24–26^. Most children likely experienced symptom onset prior to the diagnosis, and thus the demands for parental adaptation are expected to precede the diagnostic confirmation of a child’s disorder. The relative risk remained at its highest during the six months following the child’s diagnosis, and then started to decline. Such observation could feasibly result from the immediate psychological strain and reallocation of family resources associated with the onset of a child’s mental disorder or providing care and support for the child, which is then followed by a phase of psychological adaptation.^27^

In contrast to the analyses including any diagnosis of a child, results from analyses limited to specific disorders of a child differed across parents’ sex. While the temporal patterns were largely mixed among men, a time-dependent trend – markedly higher relative risk close to the time of the child’s diagnosis – was observed among women across most of the children’s diagnostic categories. This temporal pattern was generally robust after adjusting for other mental disorders of siblings or the children themselves, although the estimates did attenuate. It is important to note that in the analyses including any diagnosis of a child, only the first diagnosis among children of a parent was defined as the exposure, whereas in the disorder-specific analyses, other diagnoses could have occurred already. As such, our findings suggest that the transient elevation in the relative risk may occur in women regardless of prior comorbidity among their children, whereas for men, it is consistently observed only in relation to the first diagnosis in their family. Additionally, the relative risks associated with a child’s mental disorder were generally greater for women than for men. We hypothesize these sex differences can stem from societal and cultural expectations assigning the primary caregiving roles to women,^28^ or gendered norms that regulate experience and expression of distress.^29^

Although the results in the Finnish and the Danish data were generally consistent, we also saw differences. For instance, the transient increase in the relative risk associated with a child’s anxiety disorder was markedly greater in Denmark than in Finland. In contrast, a clear time-dependent trend associated with child’s eating disorders among women in Finland was not observed in Denmark. Although the Finnish and Danish healthcare systems are similar in many respects,^30^ we cannot rule out potential differences in diagnostic or treatment protocols, or cultural dissimilarity affecting perceptions of mental disorders. Also, the coverage of the Finnish and Danish data was different; while the Finnish data included diagnoses from both primary and secondary health care, the Danish data were limited to secondary healthcare only. Given that our sensitivity analyses using only Finnish secondary health care data suggested that the relative risk of parent receiving a mental disorder diagnosis was higher than when the primary care data was also included, the differences in coverage between Finnish and Danish data need to be considered to affect our findings.

## Limitations

The present study has limitations. First, in our study population, parents could be considered exposed only if their child received a mental disorder diagnosis in the primary or secondary healthcare. Although persons with severe mental disorders are likely to eventually present to the healthcare system, persons whose mental disorders were untreated (or were treated only within primary health care in Denmark) during the follow-up were misclassified as not having a mental disorder. Studies have shown that register-based diagnoses have good validity^31–34^, but not all disorders have been validated and the validity of diagnosis may differ in different clinical settings. Second, as discussed above, a diagnosis is not an unambiguous time zero for the onset of a child’s mental disorder. Since no information on symptoms is systematically available through registers prior to the diagnosis, we were not able to estimate temporal trends in parents’ morbidity since the child’s symptom onset. Similarly, we had no information on child’s remission or recovery, which can modify the associations with parents’ morbidity over time. Third, no specific residency criteria to ensure that parents were living in Finland or Denmark during the period when prior mental disorders were assessed was applied. This may lead to shorter exclusion period among migrant parents potentially resulting in the parent’s diagnosis during follow-up being the first one in Finland or Denmark rather than being their first mental disorder diagnosis. Fourth, our study is an observational study, and it is difficult to establish causality from its findings. Although adjustment for prior psychiatric comorbidity among children within a family and examining the time before the child’s diagnosis may capture some confounding shared among family members, we cannot rule out the possibility of unmeasured or residual confounding, which which can introduce bias in the estimated associations. Finally, the study populations were from two Nordic countries with universal access to health care and strong welfare systems, which limits the generalizability of our findings.

## Conclusions

Nationwide data from Finland and Denmark suggest that children’s mental disorders may temporarily increase their parents’ risk of mental disorders. These findings need to be further evaluated in different populations.

## Funding

The present study was funded by the European Union (ERC, MENTALNET, 101040247 to CH), the Research Council of Finland (354237 to CH; 339390 to ME), and Lundbeck Foundation (Fellowship R345-2020-1588 to OP-R). Views and opinions expressed are however those of the author(s) only and do not necessarily reflect those of the European Union or the European Research Council. Neither the European Union nor the granting authority can be held responsible for them.

## Declaration of Interest

None

## Author Contribution

All authors participated in designing the study, generating hypotheses, interpreting the data, and critically reviewing the manuscript. PB, ME, MG, CH and KK were primarily responsible for writing the manuscript. MG, RN and NM conducted the data analyses. MG, RN, NM and OP-L accessed and verified the data. All authors approved the final version of manuscript for submission.

## Data availability

Data for the present study is property of Statistics Finland, Finnish Institute of Health and Welfare, Statistics Denmark, and the Danish Health Data Authority. The data are available from these authorities, but restrictions apply.

## Notes

### Competing Interest Statement

The authors have declared no competing interest.

### Funding Statement

The present study was funded by the European Union (ERC, MENTALNET, 101040247), the Research Council of Finland (354237 to CH; 339390 to ME), and Lundbeck Foundation (Fellowship R345-2020-1588 to OP-R). Views and opinions expressed are however those of the author(s) only and do not necessarily reflect those of the European Union or the European Research Council. Neither the European Union nor the granting authority can be held responsible for them.

### Author Declarations

The Ethics Committee of the Finnish Institute for Health and Welfare approved the study plan (THL/184/6.02.01/2023:933). Data were linked with the permission of Statistics Finland (TK-53-1696-16), Statistics Denmark, and the Finnish Institute of Health and Welfare. According to Finnish and Danish law, informed consent is not required for register-based studies.

### Summary of Updates

Additional sensitivity analyses have been conducted and Supplemental file has been updated.

