## Supplement for "The Associations of Mental Disorders in Children With Parents’ Subsequent Mental Disorders: A Nationwide Cohort Study From Finland and Denmark"

**Appendix**

**Description of Study Cohorts**	3

**Description of the Comorbidity Adjustment**	4

**Supplement Table 1.** Diagnostic Categories of Mental Disorders in Finland and Denmark	5

**Supplement Table 2.** Women's Minimally and Comorbidity Adjusted Hazard Ratios and 95% Confidence Intervals for Mental Disorders in Relation to a Child’s Specific Mental Disorder According to Time Since the Child’s Diagnosis	7

**Supplement Table 3.** Men's Minimally and Comorbidity Adjusted Hazard Ratios and 95% Confidence Intervals for Mental Disorders in Relation to a Child’s Specific Mental Disorder According to Time Since the Child’s Diagnosis	9

**Supplement Table 4**. Frequencies of Persons Diagnosed Prior to Follow-up, at Risk at Start of Follow-up, Diagnosed or Otherwise Censored During Follow-up, and Exposed During Follow-up in the Sensitivity Analyses.	11

**Supplement Table 5**. Descriptives of Study Population in Sensitivity Analyses.	12

**Supplement Table 6.** Women's Minimally and Comorbidity Adjusted Hazard Ratios and 95% Confidence Intervals for Mental Disorders in Relation to a Child’s Specific Mental Disorder Before and After the Child’s Diagnosis	13

**Supplement Table 7.** Men's Minimally and Comorbidity Adjusted Hazard Ratios and 95% Confidence Intervals for Mental Disorders in Relation to a Child’s Specific Mental Disorder Before and After the Child’s Diagnosis	15

**Supplement Figure 1.** Hazard Ratios and 95% Confidence Intervals of Parents’ Mental Disorder in Relation to Any Mental Disorder of a Child Before and After the Child’s Diagnosis	17

**Supplement Figure 2.** Women's Hazard Ratios and 95% Confidence Intervals of Mental Disorder in Relation to a Child’s Specific Mental Disorder Before and After the Child’s Diagnosis	18

**Supplement Figure 3.** Men's Hazard Ratios and 95% Confidence Intervals of Mental Disorder in Relation to a Child’s Specific Mental Disorder Before and After the Child’s Diagnosis	19

**Supplement Figure 4.** Hazard Ratios and 95% Confidence Intervals of Parents’ Mental Disorder in Relation to Any Mental Disorder After the Child’s Diagnosis in Finland	20

**Supplement Figure 5.** Women's Hazard Ratios and 95% Confidence Intervals of Mental Disorder in Relation to a Child’s Specific Mental Disorder After the Child’s Diagnosis in Finland	21

**Supplement Figure 6.** Men's Hazard Ratios and 95% Confidence Intervals of Mental Disorder in Relation to a Child’s Specific Mental Disorder After the Child’s Diagnosis in Finland	22

### **Description of Study Cohorts**

**Finland**

Register data was constructed by linking the following registers: Care Register for Health Care (CRHC), the Hospital Discharge Register (HDR), and the Register of Primary Healthcare Visits (RPHV) maintained by the National Institute for Health and Welfare, and the full Finnish Population Register (FOLK) maintained by Statistics Finland. The CRHC and HDR contain information on public and private hospital admissions in Finland whereas the RPHV includes records of public and private primary healthcare visits in Finland. In the current study, HDR data from 1970 to 1993, CRHC data from 1994 onwards, and RPHV data from 2011 onwards were used. FOLK contains yearly updated administrative registers and includes basic demographic characteristics of the whole Finnish population.

**Denmark**

Register data was constructed by linking the following registers: the Central Person Register (CPR), the Danish Psychiatric Central Research Register, and the Danish Education Registers. The Central Person Register contains information on all people living in Denmark. It was established in 1968 and contains information including continually updated vital status. The Danish Psychiatric Central Research Register contains data on all psychiatric inpatient contacts since 1969, and all psychiatric outpatient and emergency contacts since 1995. The Danish Education Registers provide educational information on all individuals registered in Denmark from 1980 onwards.

### **Description of the Comorbidity Adjustment**

**Figure A** shows an example situation where the follow-up begins 1/1/2010, and the parent is unexposed. A child receives an anxiety disorder diagnosis 15/2/2013, which is adjusted as comorbid anxiety from hereon. After that, a sibling receives a developmental disorder diagnosis, which is then adjusted as comorbid developmental disorder from hereon. 7/9/2016, the child gets a depressive disorder diagnosis, and the parent is exposed from hereon. Follow-up ends at administrative censoring, parent did not have the outcome. Children’s diagnoses after exposure are not included in analyses (crossed out in the **Figure A)**.

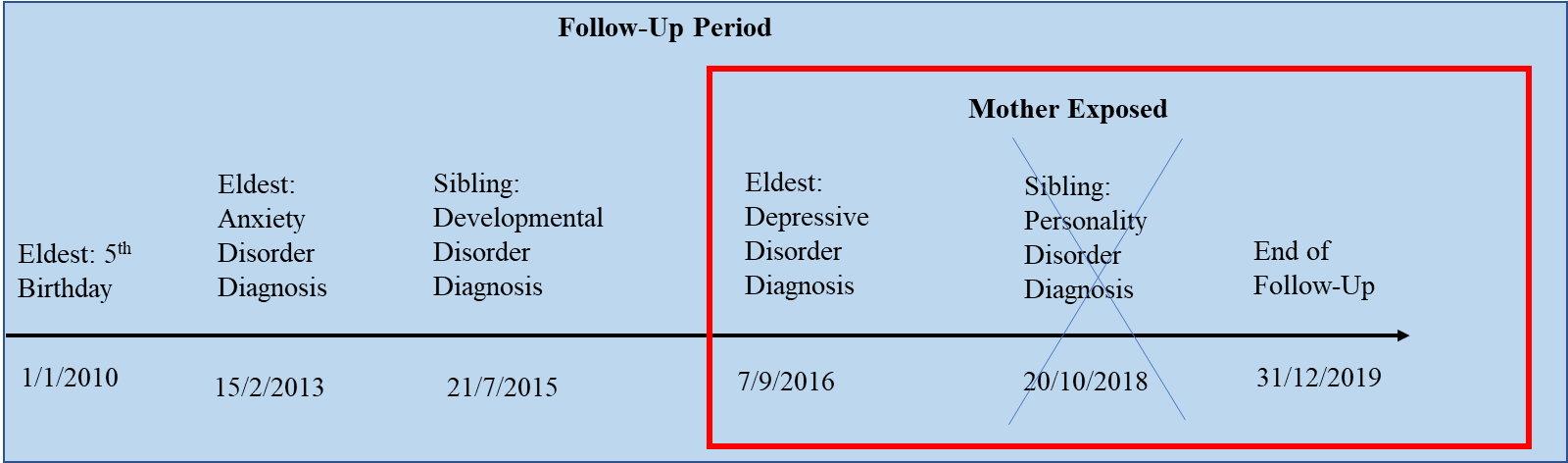

**Figure A.** Child’s depressive disorder and parents’s mental health: exposed mother

Another example situation is shown in **Figure B**; the follow-up begins 1/1/2010, the parent is unexposed. A child receives an anxiety disorder diagnosis 15/2/2013, which is adjusted as comorbid anxiety from hereon. After that, a sibling gets a developmental disorder diagnosis, which is then adjusted as comorbid developmental disorder from hereon. Sibling gets a personality disorder diagnosis, which is then adjusted as comorbid personality disorder from hereon. Follow-up ends at parent’s anxiety diagnosis. Children’s diagnoses after the end of follow-up are not included in analyses (crossed out in **Figure B**).

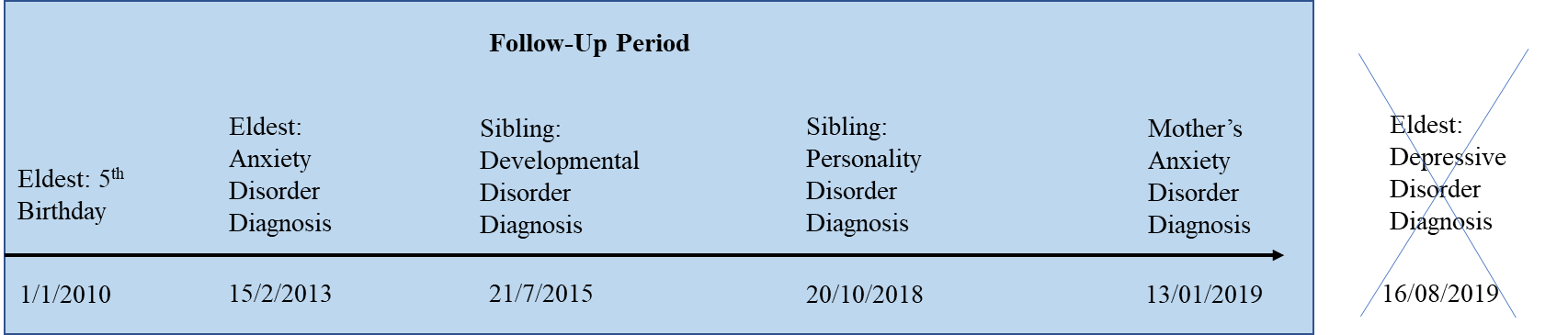

**Figure B.** Child’s depressive disorder and mother’s mental health: unexposed mother

### **Supplement Table 1.** Diagnostic Categories of Mental Disorders in Finland and Denmark

| **Mental Disorder Category** | **ICD-10** | **Abbreviation** | **Finnish Conversions** | | | **Danish Conversions** |
| --- | --- | --- | --- | --- | --- | --- |
|  |  |  | **ICD-8 Equivalents** | **ICD-9 Equivalents** | **ICPC-2 Equivalents** | **ICD-8**  **Equivalents** |
| Mental and behavioral disorders due to psychoactive substance use | F10–F19 | Substance use disorders | 291, 303, 304 | 291, 292, 303, 304, 305 | P16, P15, P17, P19 | 291.x9, 294.39, 303.x9, 303.20, 303.28, 303.90, 304.x9 |
| Schizophrenia, schizotypal and delusional disorders | F20–F29 | Psychotic disorders | 295, 297, 298.1, 298.2, 298.3, 298.9, 299 | 295, 297, 298 | P72 | 295.x9, 296.89, 297.x9, 298.29–298.99, 299.04, 299.05, 299.09, 301.83 |
| Mood (affective) disorders | F30–F39 | Mood disorders | 296, 298.0, 300.4, 301.1 | 296, 311 | P73 | 296.x9 (Excluding 296.89), 298.09, 298.19, 300.49, 301.19 |
| Neurotic, stress-related and somatoform disorders | F40–F48 | Anxiety disorders | 300.0, 300.1, 300.2, 300.3 300.5, 300.6, 300.7, 300.8, 300.9, 305, 306.8, 307, 308.4 | 300, 307.8, 309 | P79, P74, P02, P82, P75, P78 | 300.x9 (Excluding 300.49), 305.x9, 305.68, 307.99 |
| Eating Disorders | F50 | Eating Disorders | 305.60, 306.50, 306.58, 306.59 | 307.1, 307.5 | P86 | 305.60, 306.50, 306.58, 306.59 |
| Specific Personality Disorders | F60 | Personality disorders | 301.x9 (excluding 301.19), 301.80,  301.81, 301.82, 301.84 | 301 | - | 301.x9 (Excluding 301.19), 301.80, 301.81, 301.82, 301.84 |
| Intellectual disability | F70–F79 | Intellectual disabilities | 310, 311, 312, 313, 314, 315 | 317, 318, 319 | P85 | 311.xx, 312.xx, 313.xx, 314.xx, 315.xx |
| Pervasive Developmental Disorders | F84 | Developmental disorders | 299.00, 299.01, 299.02, 299.03 | 299 | - | 299.00, 299.01, 299.02, 299.03 |
| Behavioral and emotional disorders with onset in childhood and adolescence | F90–F98 | Childhood onset disorders | 306.2, 306.6, 306.7, 306.9, 308.1, 308.2 308.3 | 308.0, 307.2, 307.3, 307.6, 307.7, 307.9, 313, 314 | P81, P22, P23, P10, P12, P13, P11 | 306.x9, 308.0x |
| Any mental disorder | F00–F99 | Any mental disorder | All of the above + 290, 292, 293, 294, rest of 301, 302, 3060 3061, 3063, 3064, 3080, 309 | All of the above + 290, 293, 294, 302, 3074, 312, 316, 310, 315 | All of the above + P06, P07, P08, P09, P18, P24, P29, P70, P71, P76, P80, P98, P99 | All of the above |

### **Supplement Table 2.** Women's Minimally and Comorbidity Adjusted Hazard Ratios and 95% Confidence Intervals for Mental Disorders in Relation to a Child’s Specific Mental Disorder According to Time Since the Child’s Diagnosis

| Diagnosis | Model | Finland | | | | | Denmark | | | | |
| --- | --- | --- | --- | --- | --- | --- | --- | --- | --- | --- | --- |
|  |  | 0-6m | 6-12m | 1-1.5y | 1.5-2y | 2y+ | 0-6m | 6-12m | 1-1.5y | 1.5-2y | 2y+ |
| Any Mental Disorder | Minimally Adjusted | 2.29 (2.21, 2.38) | 1.83 (1.75, 1.91) | 1.71 (1.64, 1.79) | 1.66 (1.59, 1.74) | 1.5 (1.48, 1.53) | 2.54 (2.35, 2.75) | 2.22 (2.04, 2.42) | 1.92 (1.75, 2.11) | 1.91 (1.74, 2.11) | 1.77 (1.72, 1.83) |
| Child's Substance Use Disorder |  |  |  |  |  |  |  |  |  |  |  |
|  | Minimally Adjusted | 2.07 (1.88, 2.28) | 1.61 (1.43, 1.8) | 1.63 (1.45, 1.83) | 1.48 (1.31, 1.69) | 1.33 (1.26, 1.4) | 2.48 (1.88, 3.26) | 2.01 (1.47, 2.76) | 1.65 (1.16, 2.37) | 1.87 (1.32, 2.65) | 1.8 (1.59, 2.04) |
|  | Comorbidity Adjusted | 1.73 (1.57, 1.91) | 1.36 (1.21, 1.52) | 1.39 (1.24, 1.57) | 1.29 (1.13, 1.46) | 1.24 (1.18, 1.3) | 1.81 (1.38, 2.39) | 1.48 (1.08, 2.03) | 1.23 (0.86, 1.76) | 1.41 (1.00, 2.00) | 1.43 (1.26, 1.62) |
| Child's Psychotic Disorder |  |  |  |  |  |  |  |  |  |  |  |
|  | Minimally Adjusted | 3.71 (3.22, 4.28) | 2.05 (1.69, 2.5) | 1.86 (1.5, 2.31) | 1.83 (1.46, 2.28) | 1.51 (1.39, 1.65) | 2.48 (1.92, 3.21) | 1.76 (1.29, 2.41) | 2.05 (1.52, 2.78) | 1.78 (1.27, 2.49) | 1.82 (1.62, 2.04) |
|  | Comorbidity Adjusted | 2.61 (2.27, 3.01) | 1.46 (1.2, 1.78) | 1.34 (1.08, 1.66) | 1.33 (1.06, 1.66) | 1.18 (1.08, 1.29) | 1.76 (1.37, 2.28) | 1.26 (0.92, 1.72) | 1.47 (1.09, 1.99) | 1.29 (0.92, 1.8) | 1.38 (1.23, 1.55) |
| Child's Mood Disorder |  |  |  |  |  |  |  |  |  |  |  |
|  | Minimally Adjusted | 2.73 (2.58, 2.88) | 2.03 (1.89, 2.17) | 1.71 (1.59, 1.85) | 1.74 (1.61, 1.88) | 1.6 (1.55, 1.65) | 2.42 (2.03, 2.89) | 2.32 (1.92, 2.79) | 1.53 (1.2, 1.94) | 1.74 (1.38, 2.19) | 1.82 (1.68, 1.97) |
|  | Comorbidity Adjusted | 2.37 (2.24, 2.51) | 1.77 (1.66, 1.89) | 1.51 (1.4, 1.62) | 1.54 (1.42, 1.66) | 1.46 (1.42, 1.51) | 2.01 (1.68, 2.39) | 1.93 (1.6, 2.32) | 1.28 (1.01, 1.62) | 1.46 (1.16, 1.84) | 1.6 (1.48, 1.73) |
| Child's Anxiety Disorder |  |  |  |  |  |  |  |  |  |  |  |
|  | Minimally Adjusted | 2.31 (2.2, 2.43) | 1.86 (1.75, 1.97) | 1.55 (1.45, 1.66) | 1.66 (1.55, 1.77) | 1.51 (1.47, 1.55) | 2.86 (2.57, 3.19) | 2.1 (1.84, 2.39) | 1.95 (1.7, 2.24) | 1.82 (1.57, 2.11) | 1.78 (1.69, 1.87) |
|  | Comorbidity Adjusted | 2.06 (1.96, 2.17) | 1.66 (1.57, 1.76) | 1.39 (1.3, 1.49) | 1.49 (1.4, 1.6) | 1.39 (1.35, 1.43) | 2.54 (2.28, 2.82) | 1.86 (1.64, 2.12) | 1.74 (1.51, 1.99) | 1.63 (1.41, 1.89) | 1.63 (1.55, 1.72) |
| Child's Eating Disorder |  |  |  |  |  |  |  |  |  |  |  |
|  | Minimally Adjusted | 2.32 (2.02, 2.67) | 1.7 (1.44, 2.01) | 1.62 (1.36, 1.93) | 1.7 (1.43, 2.03) | 1.28 (1.2, 1.37) | 1.4 (0.98, 2.01) | 1.59 (1.12, 2.24) | 1.55 (1.07, 2.23) | 1.52 (1.05, 2.22) | 1.26 (1.1, 1.44) |
|  | Comorbidity Adjusted | 1.96 (1.7, 2.25) | 1.45 (1.22, 1.71) | 1.4 (1.17, 1.67) | 1.5 (1.25, 1.79) | 1.25 (1.17, 1.33) | 1.26 (0.88, 1.8) | 1.43 (1.01, 2.02) | 1.41 (0.98, 2.02) | 1.4 (0.96, 2.04) | 1.24 (1.09, 1.42) |
| Child's Personality Disorder |  |  |  |  |  |  |  |  |  |  |  |
|  | Minimally Adjusted | 1.89 (1.49, 2.4) | 1.85 (1.44, 2.37) | 1.55 (1.16, 2.07) | 1.88 (1.43, 2.48) | 1.57 (1.38, 1.78) | 2.28 (1.74, 2.97) | 2.08 (1.56, 2.77) | 2.23 (1.68, 2.97) | 1.72 (1.23, 2.41) | 1.77 (1.58, 1.98) |
|  | Comorbidity Adjusted | 1.16 (0.91, 1.47) | 1.14 (0.89, 1.47) | 0.97 (0.72, 1.29) | 1.18 (0.9, 1.56) | 1.04 (0.91, 1.18) | 1.58 (1.21, 2.06) | 1.45 (1.09, 1.94) | 1.57 (1.18, 2.09) | 1.22 (0.87, 1.71) | 1.32 (1.18, 1.49) |
| Child's Intellectual Disability |  |  |  |  |  |  |  |  |  |  |  |
|  | Minimally Adjusted | 1.9 (1.56, 2.32) | 1.43 (1.14, 1.79) | 1.58 (1.27, 1.96) | 1.52 (1.22, 1.9) | 1.23 (1.16, 1.3) | 2.06 (1.57, 2.72) | 1.69 (1.25, 2.3) | 1.41 (1, 1.98) | 1.31 (0.91, 1.87) | 1.53 (1.41, 1.67) |
|  | Comorbidity Adjusted | 1.63 (1.34, 1.99) | 1.24 (0.99, 1.56) | 1.38 (1.11, 1.72) | 1.35 (1.08, 1.68) | 1.24 (1.17, 1.31) | 1.7 (1.29, 2.24) | 1.41 (1.03, 1.91) | 1.18 (0.84, 1.65) | 1.1 (0.77, 1.57) | 1.4 (1.29, 1.53) |
| Child's Developmental Disorder |  |  |  |  |  |  |  |  |  |  |  |
|  | Minimally Adjusted | 2.82 (2.44, 3.26) | 2.11 (1.78, 2.5) | 1.89 (1.57, 2.27) | 1.84 (1.52, 2.21) | 1.43 (1.36, 1.5) | 1.81 (1.52, 2.17) | 2.14 (1.81, 2.53) | 2.29 (1.94, 2.71) | 1.93 (1.61, 2.33) | 1.61 (1.52, 1.71) |
|  | Comorbidity Adjusted | 2.2 (1.9, 2.54) | 1.67 (1.41, 1.97) | 1.51 (1.25, 1.81) | 1.48 (1.23, 1.79) | 1.37 (1.3, 1.44) | 1.54 (1.29, 1.85) | 1.83 (1.55, 2.17) | 1.98 (1.67, 2.33) | 1.68 (1.39, 2.02) | 1.52 (1.43, 1.61) |
| Child's Childhood Onset Disorder |  |  |  |  |  |  |  |  |  |  |  |
|  | Minimally Adjusted | 2.48 (2.35, 2.61) | 1.93 (1.82, 2.05) | 1.9 (1.79, 2.02) | 1.72 (1.61, 1.83) | 1.49 (1.46, 1.52) | 2.62 (2.35, 2.91) | 2.28 (2.03, 2.56) | 2.22 (1.96, 2.5) | 1.97 (1.73, 2.24) | 1.72 (1.66, 1.8) |
|  | Comorbidity Adjusted | 2.33 (2.21, 2.45) | 1.83 (1.72, 1.94) | 1.81 (1.7, 1.92) | 1.64 (1.54, 1.75) | 1.51 (1.48, 1.54) | 2.39 (2.15, 2.66) | 2.09 (1.86, 2.35) | 2.04 (1.81, 2.3) | 1.82 (1.6, 2.08) | 1.68 (1.62, 1.75) |

### **Supplement Table 3.** Men's Minimally and Comorbidity Adjusted Hazard Ratios and 95% Confidence Intervals for Mental Disorders in Relation to a Child’s Specific Mental Disorder According to Time Since the Child’s Diagnosis

| Diagnosis | Model | Finland | | | | | Denmark | | | | |
| --- | --- | --- | --- | --- | --- | --- | --- | --- | --- | --- | --- |
|  |  | 0-6m | 6-12m | 1-1.5y | 1.5-2y | 2y+ | 0-6m | 6-12m | 1-1.5y | 1.5-2y | 2y+ |
| Any Mental Disorder | Minimally Adjusted | 1.75 (1.67, 1.85) | 1.58 (1.5, 1.67) | 1.39 (1.31, 1.48) | 1.4 (1.32, 1.49) | 1.33 (1.3, 1.36) | 2.04 (1.85, 2.24) | 1.64 (1.47, 1.84) | 1.92 (1.72, 2.13) | 1.5 (1.33, 1.69) | 1.51 (1.45, 1.56) |
| Child's Substance Use Disorder |  |  |  |  |  |  |  |  |  |  |  |
|  | Minimally Adjusted | 1.72 (1.5, 1.97) | 1.63 (1.41, 1.89) | 1.43 (1.22, 1.68) | 1.62 (1.39, 1.9) | 1.37 (1.28, 1.46) | 2.59 (1.89, 3.57) | 1.01 (0.6, 1.7) | 2.07 (1.42, 3.02) | 2.36 (1.64, 3.39) | 1.97 (1.72, 2.27) |
|  | Comorbidity Adjusted | 1.51 (1.32, 1.73) | 1.45 (1.25, 1.67) | 1.28 (1.09, 1.5) | 1.46 (1.25, 1.71) | 1.3 (1.22, 1.39) | 2.05 (1.49, 2.81) | 0.8 (0.47, 1.35) | 1.65 (1.13, 2.41) | 1.89 (1.31, 2.72) | 1.64 (1.42, 1.89) |
| Child's Psychotic Disorder |  |  |  |  |  |  |  |  |  |  |  |
|  | Minimally Adjusted | 2.04 (1.6, 2.59) | 1.72 (1.32, 2.26) | 2.1 (1.63, 2.71) | 1.53 (1.13, 2.08) | 1.58 (1.42, 1.75) | 2.86 (2.17, 3.77) | 1.68 (1.16, 2.44) | 1.82 (1.25, 2.63) | 1.95 (1.34, 2.82) | 1.75 (1.53, 2.01) |
|  | Comorbidity Adjusted | 1.56 (1.22, 1.99) | 1.33 (1.02, 1.74) | 1.64 (1.27, 2.11) | 1.2 (0.88, 1.63) | 1.31 (1.18, 1.46) | 2.19 (1.66, 2.89) | 1.29 (0.89, 1.88) | 1.4 (0.97, 2.04) | 1.51 (1.04, 2.19) | 1.42 (1.23, 1.63) |
| Child's Mood Disorder |  |  |  |  |  |  |  |  |  |  |  |
|  | Minimally Adjusted | 1.78 (1.63, 1.94) | 1.69 (1.54, 1.85) | 1.4 (1.26, 1.55) | 1.64 (1.49, 1.81) | 1.44 (1.39, 1.5) | 2.12 (1.72, 2.62) | 1.79 (1.41, 2.27) | 1.97 (1.55, 2.49) | 1.3 (0.96, 1.75) | 1.63 (1.48, 1.78) |
|  | Comorbidity Adjusted | 1.6 (1.46, 1.74) | 1.52 (1.38, 1.66) | 1.26 (1.14, 1.4) | 1.49 (1.35, 1.65) | 1.35 (1.3, 1.41) | 1.85 (1.49, 2.29) | 1.56 (1.23, 1.98) | 1.72 (1.36, 2.19) | 1.14 (0.85, 1.54) | 1.48 (1.35, 1.63) |
| Child's Anxiety Disorder |  |  |  |  |  |  |  |  |  |  |  |
|  | Minimally Adjusted | 1.81 (1.68, 1.94) | 1.57 (1.45, 1.7) | 1.47 (1.35, 1.6) | 1.39 (1.27, 1.52) | 1.4 (1.35, 1.45) | 2.16 (1.88, 2.49) | 1.72 (1.46, 2.02) | 1.73 (1.46, 2.04) | 1.28 (1.05, 1.55) | 1.59 (1.5, 1.69) |
|  | Comorbidity Adjusted | 1.66 (1.54, 1.78) | 1.44 (1.33, 1.56) | 1.36 (1.25, 1.48) | 1.29 (1.18, 1.41) | 1.32 (1.27, 1.36) | 1.98 (1.72, 2.27) | 1.57 (1.34, 1.85) | 1.58 (1.34, 1.87) | 1.17 (0.96, 1.43) | 1.49 (1.41, 1.59) |
| Child's Eating Disorder |  |  |  |  |  |  |  |  |  |  |  |
|  | Minimally Adjusted | 1.61 (1.3, 1.99) | 1.51 (1.2, 1.89) | 1.19 (0.92, 1.55) | 1.4 (1.09, 1.78) | 1.17 (1.07, 1.27) | 1.79 (1.25, 2.56) | 1.26 (0.82, 1.96) | 1.98 (1.37, 2.85) | 1.08 (0.65, 1.8) | 1.26 (1.09, 1.47) |
|  | Comorbidity Adjusted | 1.42 (1.14, 1.75) | 1.34 (1.07, 1.68) | 1.07 (0.83, 1.39) | 1.27 (0.99, 1.62) | 1.15 (1.06, 1.25) | 1.65 (1.15, 2.36) | 1.17 (0.76, 1.82) | 1.85 (1.28, 2.66) | 1.02 (0.62, 1.69) | 1.25 (1.08, 1.46) |
| Child's Personality Disorder |  |  |  |  |  |  |  |  |  |  |  |
|  | Minimally Adjusted | 1.41 (1, 1.98) | 2.23 (1.68, 2.96) | 1.55 (1.08, 2.22) | 1.87 (1.33, 2.64) | 1.21 (1.02, 1.44) | 1.89 (1.35, 2.65) | 1.82 (1.28, 2.58) | 2.14 (1.53, 2.99) | 1.34 (0.86, 2.07) | 1.51 (1.31, 1.74) |
|  | Comorbidity Adjusted | 0.97 (0.69, 1.37) | 1.55 (1.17, 2.06) | 1.08 (0.76, 1.55) | 1.31 (0.93, 1.85) | 0.89 (0.75, 1.06) | 1.38 (0.99, 1.94) | 1.33 (0.94, 1.9) | 1.58 (1.13, 2.21) | 0.99 (0.64, 1.54) | 1.18 (1.02, 1.36) |
| Child's Intellectual Disability |  |  |  |  |  |  |  |  |  |  |  |
|  | Minimally Adjusted | 1.65 (1.26, 2.14) | 1.67 (1.29, 2.17) | 1.39 (1.04, 1.86) | 1.03 (0.74, 1.44) | 1.25 (1.16, 1.34) | 1.58 (1.09, 2.29) | 2.07 (1.49, 2.87) | 1.54 (1.05, 2.26) | 1.51 (1.02, 2.24) | 1.48 (1.33, 1.63) |
|  | Comorbidity Adjusted | 1.48 (1.14, 1.93) | 1.51 (1.16, 1.97) | 1.27 (0.95, 1.7) | 0.95 (0.68, 1.33) | 1.27 (1.18, 1.36) | 1.38 (0.96, 2.01) | 1.82 (1.31, 2.52) | 1.36 (0.93, 2) | 1.34 (0.91, 1.99) | 1.4 (1.26, 1.55) |
| Child's Developmental Disorder |  |  |  |  |  |  |  |  |  |  |  |
|  | Minimally Adjusted | 1.65 (1.31, 2.08) | 1.25 (0.96, 1.63) | 1.78 (1.42, 2.24) | 1.52 (1.19, 1.95) | 1.36 (1.28, 1.45) | 1.82 (1.49, 2.23) | 1.35 (1.06, 1.71) | 1.51 (1.2, 1.9) | 1.6 (1.28, 2.01) | 1.27 (1.18, 1.36) |
|  | Comorbidity Adjusted | 1.39 (1.1, 1.75) | 1.06 (0.81, 1.39) | 1.52 (1.21, 1.92) | 1.32 (1.03, 1.69) | 1.32 (1.24, 1.4) | 1.61 (1.31, 1.97) | 1.19 (0.94, 1.52) | 1.35 (1.07, 1.7) | 1.44 (1.14, 1.81) | 1.21 (1.13, 1.3) |
| Child's Childhood Onset Disorder |  |  |  |  |  |  |  |  |  |  |  |
|  | Minimally Adjusted | 1.8 (1.67, 1.94) | 1.71 (1.59, 1.85) | 1.52 (1.4, 1.65) | 1.49 (1.37, 1.62) | 1.34 (1.3, 1.37) | 2.15 (1.88, 2.46) | 1.84 (1.59, 2.13) | 2.01 (1.74, 2.33) | 1.84 (1.58, 2.14) | 1.46 (1.39, 1.53) |
|  | Comorbidity Adjusted | 1.72 (1.6, 1.85) | 1.65 (1.52, 1.78) | 1.47 (1.35, 1.6) | 1.45 (1.33, 1.57) | 1.35 (1.32, 1.39) | 2.02 (1.76, 2.31) | 1.73 (1.49, 2.01) | 1.9 (1.65, 2.2) | 1.75 (1.5, 2.03) | 1.44 (1.37, 1.51) |

### **Supplement Table 4**. Frequencies of Persons Diagnosed Prior to Follow-up, at Risk at Start of Follow-up, Diagnosed or Otherwise Censored During Follow-up, and Exposed During Follow-up in the Sensitivity Analyses.

|  |  | Finland | | Denmark | |
| --- | --- | --- | --- | --- | --- |
|  |  | Women | Men | Women | Men |
| Mental Disorder Diagnosis Prior to Follow-up |  |  |  |  |  |
|  | Early Follow-up Start | 17893 | 21768 | 17883 | 12141 |
|  | Late Follow-up Start | 28696 | 29331 | 27641 | 18625 |
| Persons at Risk at Start of Follow-up |  |  |  |  |  |
|  | Early Follow-up Start | 394700 | 381683 | 444013 | 441125 |
|  | Late Follow-up Start | 381476 | 371345 | 425104 | 424032 |
| New Cases During Follow-up |  |  |  |  |  |
|  | Early Follow-up Start | 107203 | 68787 | 54007 | 40705 |
|  | Late Follow-up Start | 96400 | 61224 | 44249 | 34221 |
| Emigrated During Follow-up |  |  |  |  |  |
|  | Early Follow-up Start | 7823 | 7735 | 24267 | 29914 |
|  | Late Follow-up Start | 5693 | 5986 | 15495 | 20316 |
| Died During Follow-up |  |  |  |  |  |
|  | Early Follow-up Start | 3167 | 8098 | 6206 | 11797 |
|  | Late Follow-up Start | 2876 | 7072 | 5827 | 10786 |
| Exposed During Follow-up |  |  |  |  |  |
|  | Any Mental Disorder | 160904 | 160752 | 97233 | 97848 |
|  | Substance Use Disorders | 17585 | 17453 | 6996 | 6818 |
|  | Psychotic Disorders | 4995 | 5088 | 9184 | 9050 |
|  | Mood Disorders | 45697 | 46877 | 21426 | 21426 |
|  | Anxiety Disorders | 66856 | 68420 | 45093 | 45648 |
|  | Eating Disorders | 8846 | 8969 | 8963 | 8932 |
|  | Personality Disorders | 3194 | 3379 | 8557 | 8606 |
|  | Intellectual Disabilities | 6366 | 6429 | 6685 | 6702 |
|  | Developmental Disorders | 8288 | 8573 | 22418 | 22892 |
|  | Childhood Onset Disorders | 67974 | 69553 | 40533 | 41307 |

### **Supplement Table 5**. Descriptives of Study Population in Sensitivity Analyses.

|  | | |  | Finland |  |  |  | Denmark |  |  |  |
| --- | --- | --- | --- | --- | --- | --- | --- | --- | --- | --- | --- |
|  | | |  | Women |  | Men |  | Women |  | Men |  |
|  | | |  | Follow-up Started at 1 | Follow-up Started at 5 | Follow-up Started at 1 | Follow-up Started at 5 | Follow-up Started at 1 | Follow-up Started at 5 | Follow-up Started at 1 | Follow-up Started at 5 |
|  | | |  | *Median (IQR)* | *Median (IQR)* | *Median (IQR)* | *Median (IQR)* | *Median (IQR)* | *Median (IQR)* | *Median (IQR)* | *Median (IQR)* |
| Parent's Birth Year, *N (%)* | | |  |  |  |  |  |  |  |  |  |
|  | <1950 | | | 735 (0.2%) | 726 (0.2%) | 4087 (1.1%) | 3964 (1.1%) | 387 (<0.1%) | 370 (<0.1%) | 4262 (1.0%) | 4043 (1.0%) |
|  | 1951-1960 | | | 29677 (7.5%) | 29086 (7.6%) | 50925 (13%) | 49969 (13%) | 26339 (5.9%) | 25378 (6.0%) | 54635 (12%) | 52760 (12%) |
|  | 1961-1970 | | | 188805 (48%) | 184661 (48%) | 195078 (51%) | 191283 (52%) | 215224 (48%) | 207893 (49%) | 232405 (53%) | 224633 (53%) |
|  | 1971-1980 | | | 152577 (39%) | 146342 (38%) | 116492 (31%) | 112384 (30%) | 183941 (41%) | 175285 (41%) | 139955 (32%) | 133762 (32%) |
|  | >1980 | | | 22906 (5.8%) | 20661 (5.4%) | 15101 (4.0%) | 13745 (3.7%) | 18122 (4.1%) | 16178 (3.8%) | 9868 (2.2%) | 8834 (2.1%) |
| Parent's Age at Start of Follow-up | | |  | 28 (24, 31) | 32 (28, 35) | 29 (26, 33) | 33 (30, 37) | 27 (25, 30) | 31 (29, 34) | 29 (26, 33) | 33 (30, 37) |
| Parent's Age at End of Follow-up | | |  | 47 (42, 53) | 48 (42, 53) | 50 (44, 55) | 50 (45, 55) | 50 (44, 55) | 50 (45, 55) | 52 (46, 57) | 52 (47, 57) |
| Parent's Education at Start of Follow-up, *N (%)* | | |  |  |  |  |  |  |  |  |  |
|  | | Lower secondary or less | | 58007 (15%) | 46791 (12%) | 71952 (19%) | 61495 (17%) | 96402 (22%) | 83887 (20%) | 96529 (22%) | 85948 (20%) |
|  | | Upper secondary | | 157774 (40%) | 146351 (38%) | 184977 (48%) | 173620 (47%) | 227832 (52%) | 214542 (51%) | 252070 (58%) | 242285 (58%) |
|  | | Post-secondary or tertiary | | 178919 (45%) | 188334 (49%) | 124754 (33%) | 136230 (37%) | 112635 (26%) | 123802 (29%) | 84239 (19%) | 91895 (22%) |
| Child's Age at Substance Use Disorder Diagnosis | | |  |  | 17 (15, 20) |  | 17 (15, 20) |  | 19 (17, 22) |  | 19 (17, 22) |
| Child's Age at Psychotic Disorder Diagnosis | | |  |  | 17 (15, 20) |  | 17 (15, 20) |  | 19 (16, 21) |  | 18 (16, 21) |
| Child's Age at Mood Disorder Diagnosis | | |  |  | 16 (14, 19) |  | 16 (14, 19) |  | 18 (16, 21) |  | 18 (16, 21) |
| Child's Age at Anxiety Disorder Diagnosis | | |  |  | 16 (13, 19) |  | 16 (13, 19) |  | 16 (14, 19) |  | 16 (14, 19) |
| Child's Age at Eating Disorder Diagnosis | | |  |  | 15 (13, 17) |  | 15 (13, 17) |  | 16 (14, 19) |  | 16 (14, 19) |
| Child's Age at Personality Disorder Diagnosis | | |  |  | 19 (18, 21) |  | 19 (18, 21) |  | 19 (17, 22) |  | 19 (17, 22) |
| Child's Age at Intellectual Disability Diagnosis | | |  | 6 (4, 12) |  | 6 (4, 12) |  | 11 (6, 16) |  | 12 (7, 16) |  |
| Child's Age at Developmental Disorder Diagnosis | | |  | 8 (5, 12) |  | 8 (5, 12) |  | 12 (8, 15) |  | 12 (8, 15) |  |
| Child's Age at Childhood Onset Disorder Diagnosis | | |  | 8 (5, 12) |  | 8 (5, 12) |  | 12 (8, 15) |  | 12 (8, 15) |  |
| Child's Age at First Mental Disorder Diagnosis | | |  | 11 (5, 16) |  | 11 (5, 16) |  | 15 (10, 18) |  | 15 (10, 18) |  |

### **Supplement Table 6.** Women's Minimally and Comorbidity Adjusted Hazard Ratios and 95% Confidence Intervals for Mental Disorders in Relation to a Child’s Specific Mental Disorder Before and After the Child’s Diagnosis

| Diagnosis | Model | Finland | | | | | | | Denmark | | | | | | |
| --- | --- | --- | --- | --- | --- | --- | --- | --- | --- | --- | --- | --- | --- | --- | --- |
|  |  | Before |  | After |  |  |  |  | Before |  | After |  |  |  |  |
|  |  | 12-6m | 6-0m | 0-6m | 6-12m | 1-1.5y | 1.5-2y | 2y+ | 12-6m | 6-0m | 0-6m | 6-12m | 1-1.5y | 1.5-2y | 2y+ |
| Any Mental Disorder | Minimally Adjusted | 1.8 (1.73, 1.88) | 2.26 (2.18, 2.35) | 2.42 (2.33, 2.52) | 1.91 (1.83, 1.99) | 1.8 (1.72, 1.88) | 1.74 (1.66, 1.82) | 1.58 (1.55, 1.6) | 1.99 (1.83, 2.17) | 2.65 (2.45, 2.85) | 2.76 (2.56, 2.98) | 2.29 (2.11, 2.5) | 1.99 (1.82, 2.19) | 1.97 (1.79, 2.17) | 1.84 (1.78, 1.9) |
| Child's Substance Use Disorder |  |  |  |  |  |  |  |  |  |  |  |  |  |  |  |
|  | Minimally Adjusted | 1.66 (1.49, 1.84) | 2.03 (1.85, 2.24) | 2.09 (1.9, 2.31) | 1.61 (1.44, 1.81) | 1.65 (1.47, 1.86) | 1.48 (1.3, 1.68) | 1.34 (1.27, 1.41) | 2.29 (1.74,  3) | 1.87 (1.38, 2.55) | 2.43 (1.84, 3.21) | 2.07 (1.51, 2.82) | 1.65 (1.15, 2.35) | 1.88 (1.33, 2.67) | 1.8 (1.59, 2.04) |
|  | Comorbidity Adjusted | 1.35 (1.21, 1.51) | 1.68 (1.53, 1.85) | 1.75 (1.59, 1.93) | 1.37 (1.22, 1.53) | 1.42 (1.27, 1.6) | 1.29 (1.14, 1.47) | 1.26 (1.2, 1.33) | 1.62 (1.24, 2.13) | 1.34 (0.98, 1.82) | 1.75 (1.33, 2.31) | 1.5 (1.1, 2.05) | 1.21 (0.84, 1.73) | 1.4 (0.99, 1.98) | 1.41 (1.25, 1.6) |
| Child's Psychotic Disorder |  |  |  |  |  |  |  |  |  |  |  |  |  |  |  |
|  | Minimally Adjusted | 1.58 (1.28, 1.96) | 2.38  (2,  2.83) | 3.76 (3.26, 4.33) | 2.06 (1.69, 2.5) | 1.86 (1.5,  2.3) | 1.84 (1.47, 2.3) | 1.51 (1.39, 1.65) | 2.05 (1.58, 2.68) | 2.26 (1.75, 2.93) | 2.49 (1.93, 3.21) | 1.76 (1.29, 2.41) | 2.04 (1.51, 2.77) | 1.74 (1.23, 2.45) | 1.83 (1.62, 2.05) |
|  | Comorbidity Adjusted | 1.06 (0.86, 1.31) | 1.62 (1.36, 1.93) | 2.58 (2.24, 2.98) | 1.43 (1.18, 1.75) | 1.31 (1.06, 1.62) | 1.31 (1.05, 1.64) | 1.18 (1.08, 1.28) | 1.41 (1.08, 1.84) | 1.57 (1.21, 2.03) | 1.73 (1.34, 2.24) | 1.23 (0.9, 1.69) | 1.44 (1.06, 1.95) | 1.23 (0.88, 1.74) | 1.37 (1.21, 1.54) |
| Child's Mood Disorder |  |  |  |  |  |  |  |  |  |  |  |  |  |  |  |
|  | Minimally Adjusted | 1.81 (1.7, 1.94) | 2.43 (2.29, 2.57) | 2.81 (2.65, 2.97) | 2.07 (1.93, 2.21) | 1.76 (1.63, 1.9) | 1.78 (1.65, 1.93) | 1.64 (1.59, 1.69) | 2.16 (1.81, 2.57) | 2.41 (2.03, 2.86) | 2.42 (2.03, 2.89) | 2.33 (1.94, 2.81) | 1.55 (1.22, 1.96) | 1.74 (1.38, 2.19) | 1.83 (1.69, 1.98) |
|  | Comorbidity Adjusted | 1.54 (1.43, 1.64) | 2.07 (1.95, 2.2) | 2.41 (2.28, 2.55) | 1.79 (1.67, 1.91) | 1.53 (1.42, 1.65) | 1.56 (1.44, 1.68) | 1.49 (1.45, 1.54) | 1.74 (1.46, 2.08) | 1.96 (1.65, 2.33) | 1.98 (1.66, 2.37) | 1.92 (1.59, 2.31) | 1.28 (1.01, 1.63) | 1.44 (1.15, 1.82) | 1.6 (1.48, 1.74) |
| Child's Anxiety Disorder |  |  |  |  |  |  |  |  |  |  |  |  |  |  |  |
|  | Minimally Adjusted | 1.85 (1.75, 1.96) | 2.48 (2.36, 2.6) | 2.45 (2.33, 2.57) | 1.92 (1.81, 2.03) | 1.61 (1.5, 1.72) | 1.72 (1.61, 1.84) | 1.57 (1.52, 1.61) | 1.95 (1.72, 2.21) | 3.37 (3.06, 3.71) | 3.24 (2.92, 3.58) | 2.16 (1.9, 2.45) | 1.99 (1.74, 2.29) | 1.84 (1.58, 2.13) | 1.81 (1.72, 1.91) |
|  | Comorbidity Adjusted | 1.64 (1.55, 1.74) | 2.21 (2.1, 2.32) | 2.18 (2.08, 2.3) | 1.72 (1.62, 1.82) | 1.44 (1.35, 1.54) | 1.55 (1.45, 1.66) | 1.44 (1.4, 1.49) | 1.72 (1.52, 1.95) | 2.99 (2.72, 3.3) | 2.88 (2.6, 3.19) | 1.92 (1.69, 2.19) | 1.79 (1.56, 2.05) | 1.65 (1.42, 1.91) | 1.67 (1.59, 1.76) |
| Child's Eating Disorder |  |  |  |  |  |  |  |  |  |  |  |  |  |  |  |
|  | Minimally Adjusted | 1.39 (1.16, 1.67) | 1.89 (1.62, 2.21) | 2.33 (2.02, 2.68) | 1.72 (1.45, 2.03) | 1.63 (1.37, 1.94) | 1.69 (1.42, 2.02) | 1.28 (1.2, 1.37) | 1.56 (1.12, 2.16) | 1.38 (0.97, 1.96) | 1.5 (1.06, 2.12) | 1.59 (1.12, 2.25) | 1.54 (1.07, 2.22) | 1.54 (1.05, 2.24) | 1.26 (1.11, 1.45) |
|  | Comorbidity Adjusted | 1.14 (0.95, 1.37) | 1.58 (1.35, 1.84) | 1.96 (1.71, 2.26) | 1.47 (1.24, 1.73) | 1.42 (1.19, 1.69) | 1.5 (1.25, 1.79) | 1.28 (1.2, 1.37) | 1.37 (0.99, 1.9) | 1.22 (0.86, 1.73) | 1.34 (0.95, 1.9) | 1.43 (1.01, 2.03) | 1.4 (0.98, 2.02) | 1.42 (0.97, 2.07) | 1.26 (1.1, 1.44) |
| Child's Personality Disorder |  |  |  |  |  |  |  |  |  |  |  |  |  |  |  |
|  | Minimally Adjusted | 1.91 (1.52, 2.41) | 1.92 (1.53, 2.42) | 1.89 (1.49, 2.39) | 1.85 (1.44, 2.37) | 1.51 (1.12, 2.02) | 1.89 (1.44, 2.49) | 1.57 (1.38, 1.78) | 2 (1.52, 2.62) | 2.67 (2.1, 3.39) | 2.29 (1.75, 2.99) | 2.09 (1.57, 2.78) | 2.23 (1.68, 2.97) | 1.74 (1.24, 2.44) | 1.77 (1.58, 1.98) |
|  | Comorbidity Adjusted | 1.11 (0.88, 1.4) | 1.13 (0.9, 1.42) | 1.12 (0.89, 1.43) | 1.11 (0.86, 1.43) | 0.89 (0.66, 1.2) | 1.16 (0.88, 1.53) | 1.02 (0.89, 1.15) | 1.33 (1.01, 1.75) | 1.79 (1.41, 2.28) | 1.55 (1.18, 2.02) | 1.42 (1.07, 1.9) | 1.53 (1.15, 2.04) | 1.21 (0.86, 1.69) | 1.3 (1.16, 1.46) |
| Child's Intellectual Disability |  |  |  |  |  |  |  |  |  |  |  |  |  |  |  |
|  | Minimally Adjusted | 1.91 (1.56, 2.34) | 1.57 (1.26, 1.96) | 1.89 (1.55, 2.31) | 1.42 (1.13, 1.78) | 1.6 (1.29, 1.98) | 1.54 (1.23, 1.92) | 1.23 (1.17, 1.3) | 1.5 (1.09, 2.06) | 2.12 (1.62, 2.77) | 2.05 (1.56, 2.71) | 1.68 (1.23, 2.29) | 1.42 (1.01, 2) | 1.33 (0.93, 1.91) | 1.55 (1.42, 1.69) |
|  | Comorbidity Adjusted | 1.61 (1.31, 1.97) | 1.32 (1.06, 1.65) | 1.63 (1.34, 1.99) | 1.24 (0.98, 1.55) | 1.41 (1.14, 1.75) | 1.38 (1.1, 1.72) | 1.29 (1.22, 1.36) | 1.21 (0.88, 1.66) | 1.72 (1.31, 2.25) | 1.69 (1.28, 2.23) | 1.39 (1.02, 1.89) | 1.19 (0.84, 1.67) | 1.12 (0.78, 1.6) | 1.43 (1.32, 1.56) |
| Child's Developmental Disorder |  |  |  |  |  |  |  |  |  |  |  |  |  |  |  |
|  | Minimally Adjusted | 2.19 (1.85, 2.58) | 2.3 (1.96, 2.7) | 2.82 (2.44, 3.26) | 2.11 (1.78, 2.5) | 1.9 (1.58, 2.28) | 1.85 (1.54, 2.23) | 1.43 (1.36, 1.5) | 1.75 (1.46, 2.09) | 1.79 (1.5, 2.13) | 1.81 (1.51, 2.16) | 2.13 (1.8, 2.52) | 2.29 (1.94, 2.7) | 1.95 (1.62, 2.35) | 1.61 (1.52, 1.71) |
|  | Comorbidity Adjusted | 1.64 (1.39, 1.93) | 1.74 (1.48, 2.04) | 2.15 (1.86, 2.49) | 1.64 (1.38, 1.94) | 1.5 (1.25, 1.8) | 1.49 (1.23, 1.79) | 1.39 (1.33, 1.46) | 1.45 (1.21, 1.73) | 1.49 (1.25, 1.78) | 1.52 (1.27, 1.82) | 1.81 (1.52, 2.14) | 1.96 (1.66, 2.31) | 1.68 (1.4, 2.02) | 1.53 (1.45, 1.62) |
| Child's Childhood Onset Disorder |  |  |  |  |  |  |  |  |  |  |  |  |  |  |  |
|  | Minimally Adjusted | 2.1 (1.99, 2.23) | 2.56 (2.43, 2.7) | 2.57 (2.44, 2.71) | 1.98 (1.87, 2.1) | 1.95 (1.84, 2.07) | 1.76 (1.65, 1.88) | 1.52 (1.49, 1.55) | 2.29 (2.05, 2.56) | 2.21 (1.97, 2.48) | 2.66 (2.39, 2.96) | 2.33 (2.07, 2.62) | 2.25 (2, 2.54) | 2.01 (1.76, 2.29) | 1.75 (1.68, 1.83) |
|  | Comorbidity Adjusted | 1.96 (1.85, 2.07) | 2.4 (2.27, 2.52) | 2.42 (2.3, 2.55) | 1.88 (1.77, 1.99) | 1.86 (1.76, 1.98) | 1.69 (1.59, 1.8) | 1.57 (1.54, 1.61) | 2.05 (1.84, 2.3) | 1.99 (1.77, 2.23) | 2.41 (2.17, 2.68) | 2.13 (1.89, 2.39) | 2.07 (1.83, 2.33) | 1.85 (1.63, 2.11) | 1.73 (1.66, 1.8) |

### **Supplement Table 7.** Men's Minimally and Comorbidity Adjusted Hazard Ratios and 95% Confidence Intervals for Mental Disorders in Relation to a Child’s Specific Mental Disorder Before and After the Child’s Diagnosis

| Diagnosis | Model | Finland | | | | | | | Denmark | | | | | | |
| --- | --- | --- | --- | --- | --- | --- | --- | --- | --- | --- | --- | --- | --- | --- | --- |
|  |  | Before |  | After |  |  |  |  | Before |  | After |  |  |  |  |
|  |  | 12-6m | 6-0m | 0-6m | 6-12m | 1-1.5y | 1.5-2y | 2y+ | 12-6m | 6-0m | 0-6m | 6-12m | 1-1.5y | 1.5-2y | 2y+ |
| Any Mental Disorder | Minimally Adjusted | 1.7 (1.61, 1.79) | 1.82 (1.72, 1.91) | 1.82 (1.72, 1.91) | 1.64 (1.55, 1.73) | 1.43 (1.35, 1.52) | 1.45 (1.37, 1.54) | 1.37 (1.35, 1.4) | 1.68 (1.52, 1.87) | 2.21 (2.02, 2.43) | 2.14 (1.95, 2.36) | 1.68 (1.5, 1.87) | 1.94 (1.74, 2.15) | 1.53 (1.35, 1.72) | 1.54 (1.48, 1.6) |
| Child's Substance Use Disorder |  |  |  |  |  |  |  |  |  |  |  |  |  |  |  |
|  | Minimally Adjusted | 1.66 (1.49, 1.84) | 2.03 (1.85, 2.24) | 2.09 (1.9, 2.31) | 1.61 (1.44, 1.81) | 1.65 (1.47, 1.86) | 1.48 (1.3, 1.68) | 1.34 (1.27, 1.41) | 2.29 (1.74, 3) | 1.87 (1.38, 2.55) | 2.43 (1.84, 3.21) | 2.07 (1.51, 2.82) | 1.65 (1.15, 2.35) | 1.88 (1.33, 2.67) | 1.8 (1.59, 2.04) |
|  | Comorbidity Adjusted | 1.35 (1.21, 1.51) | 1.68 (1.53, 1.85) | 1.75 (1.59, 1.93) | 1.37 (1.22, 1.53) | 1.42 (1.27, 1.6) | 1.29 (1.14, 1.47) | 1.26 (1.2, 1.33) | 1.62 (1.24, 2.13) | 1.34 (0.98, 1.82) | 1.75 (1.33, 2.31) | 1.5 (1.1, 2.05) | 1.21 (0.84, 1.73) | 1.4 (0.99, 1.98) | 1.41 (1.25, 1.6) |
| Child's Psychotic Disorder |  |  |  |  |  |  |  |  |  |  |  |  |  |  |  |
|  | Minimally Adjusted | 1.58 (1.28, 1.96) | 2.38  (2,  2.83) | 3.76 (3.26, 4.33) | 2.06 (1.69, 2.5) | 1.86 (1.5, 2.3) | 1.84 (1.47, 2.3) | 1.51 (1.39, 1.65) | 2.05 (1.58, 2.68) | 2.26 (1.75, 2.93) | 2.49 (1.93, 3.21) | 1.76 (1.29, 2.41) | 2.04 (1.51, 2.77) | 1.74 (1.23, 2.45) | 1.83 (1.62, 2.05) |
|  | Comorbidity Adjusted | 1.06 (0.86, 1.31) | 1.62 (1.36, 1.93) | 2.58 (2.24, 2.98) | 1.43 (1.18, 1.75) | 1.31 (1.06, 1.62) | 1.31 (1.05, 1.64) | 1.18 (1.08, 1.28) | 1.41 (1.08, 1.84) | 1.57 (1.21, 2.03) | 1.73 (1.34, 2.24) | 1.23 (0.9, 1.69) | 1.44 (1.06, 1.95) | 1.23 (0.88, 1.74) | 1.37 (1.21, 1.54) |
| Child's Mood Disorder |  |  |  |  |  |  |  |  |  |  |  |  |  |  |  |
|  | Minimally Adjusted | 1.81 (1.7, 1.94) | 2.43 (2.29, 2.57) | 2.81 (2.65, 2.97) | 2.07 (1.93, 2.21) | 1.76 (1.63, 1.9) | 1.78 (1.65, 1.93) | 1.64 (1.59, 1.69) | 2.16 (1.81, 2.57) | 2.41 (2.03, 2.86) | 2.42 (2.03, 2.89) | 2.33 (1.94, 2.81) | 1.55 (1.22, 1.96) | 1.74 (1.38, 2.19) | 1.83 (1.69, 1.98) |
|  | Comorbidity Adjusted | 1.54 (1.43, 1.64) | 2.07 (1.95, 2.2) | 2.41 (2.28, 2.55) | 1.79 (1.67, 1.91) | 1.53 (1.42, 1.65) | 1.56 (1.44, 1.68) | 1.49 (1.45, 1.54) | 1.74 (1.46, 2.08) | 1.96 (1.65, 2.33) | 1.98 (1.66, 2.37) | 1.92 (1.59, 2.31) | 1.28 (1.01, 1.63) | 1.44 (1.15, 1.82) | 1.6 (1.48, 1.74) |
| Child's Anxiety Disorder |  |  |  |  |  |  |  |  |  |  |  |  |  |  |  |
|  | Minimally Adjusted | 1.85 (1.75, 1.96) | 2.48 (2.36, 2.6) | 2.45 (2.33, 2.57) | 1.92 (1.81, 2.03) | 1.61 (1.5, 1.72) | 1.72 (1.61, 1.84) | 1.57 (1.52, 1.61) | 1.95 (1.72, 2.21) | 3.37 (3.06, 3.71) | 3.24 (2.92, 3.58) | 2.16 (1.9, 2.45) | 1.99 (1.74, 2.29) | 1.84 (1.58, 2.13) | 1.81 (1.72, 1.91) |
|  | Comorbidity Adjusted | 1.64 (1.55, 1.74) | 2.21 (2.1, 2.32) | 2.18 (2.08, 2.3) | 1.72 (1.62, 1.82) | 1.44 (1.35, 1.54) | 1.55 (1.45, 1.66) | 1.44 (1.4, 1.49) | 1.72 (1.52, 1.95) | 2.99 (2.72, 3.3) | 2.88 (2.6, 3.19) | 1.92 (1.69, 2.19) | 1.79 (1.56, 2.05) | 1.65 (1.42, 1.91) | 1.67 (1.59, 1.76) |
| Child's Eating Disorder |  |  |  |  |  |  |  |  |  |  |  |  |  |  |  |
|  | Minimally Adjusted | 1.39 (1.16, 1.67) | 1.89 (1.62, 2.21) | 2.33 (2.02, 2.68) | 1.72 (1.45, 2.03) | 1.63 (1.37, 1.94) | 1.69 (1.42, 2.02) | 1.28 (1.2, 1.37) | 1.56 (1.12, 2.16) | 1.38 (0.97, 1.96) | 1.5 (1.06, 2.12) | 1.59 (1.12, 2.25) | 1.54 (1.07, 2.22) | 1.54 (1.05, 2.24) | 1.26 (1.11, 1.45) |
|  | Comorbidity Adjusted | 1.14 (0.95, 1.37) | 1.58 (1.35, 1.84) | 1.96 (1.71, 2.26) | 1.47 (1.24, 1.73) | 1.42 (1.19, 1.69) | 1.5 (1.25, 1.79) | 1.28 (1.2, 1.37) | 1.37 (0.99, 1.9) | 1.22 (0.86, 1.73) | 1.34 (0.95, 1.9) | 1.43 (1.01, 2.03) | 1.4 (0.98, 2.02) | 1.42 (0.97, 2.07) | 1.26 (1.1, 1.44) |
| Child's Personality Disorder |  |  |  |  |  |  |  |  |  |  |  |  |  |  |  |
|  | Minimally Adjusted | 1.91 (1.52, 2.41) | 1.92 (1.53, 2.42) | 1.89 (1.49, 2.39) | 1.85 (1.44, 2.37) | 1.51 (1.12, 2.02) | 1.89 (1.44, 2.49) | 1.57 (1.38, 1.78) | 2 (1.52, 2.62) | 2.67 (2.1, 3.39) | 2.29 (1.75, 2.99) | 2.09 (1.57, 2.78) | 2.23 (1.68, 2.97) | 1.74 (1.24, 2.44) | 1.77 (1.58, 1.98) |
|  | Comorbidity Adjusted | 1.11 (0.88, 1.4) | 1.13 (0.9, 1.42) | 1.12 (0.89, 1.43) | 1.11 (0.86, 1.43) | 0.89 (0.66, 1.2) | 1.16 (0.88, 1.53) | 1.02 (0.89, 1.15) | 1.33 (1.01, 1.75) | 1.79 (1.41, 2.28) | 1.55 (1.18, 2.02) | 1.42 (1.07, 1.9) | 1.53 (1.15, 2.04) | 1.21 (0.86, 1.69) | 1.3 (1.16, 1.46) |
| Child's Intellectual Disability |  |  |  |  |  |  |  |  |  |  |  |  |  |  |  |
|  | Minimally Adjusted | 1.91 (1.56, 2.34) | 1.57 (1.26, 1.96) | 1.89 (1.55, 2.31) | 1.42 (1.13, 1.78) | 1.6 (1.29, 1.98) | 1.54 (1.23, 1.92) | 1.23 (1.17, 1.3) | 1.5 (1.09, 2.06) | 2.12 (1.62, 2.77) | 2.05 (1.56, 2.71) | 1.68 (1.23, 2.29) | 1.42 (1.01, 2) | 1.33 (0.93, 1.91) | 1.55 (1.42, 1.69) |
|  | Comorbidity Adjusted | 1.61 (1.31, 1.97) | 1.32 (1.06, 1.65) | 1.63 (1.34, 1.99) | 1.24 (0.98, 1.55) | 1.41 (1.14, 1.75) | 1.38 (1.1, 1.72) | 1.29 (1.22, 1.36) | 1.21 (0.88, 1.66) | 1.72 (1.31, 2.25) | 1.69 (1.28, 2.23) | 1.39 (1.02, 1.89) | 1.19 (0.84, 1.67) | 1.12 (0.78, 1.6) | 1.43 (1.32, 1.56) |
| Child's Developmental Disorder |  |  |  |  |  |  |  |  |  |  |  |  |  |  |  |
|  | Minimally Adjusted | 2.19 (1.85, 2.58) | 2.3 (1.96, 2.7) | 2.82 (2.44, 3.26) | 2.11 (1.78, 2.5) | 1.9 (1.58, 2.28) | 1.85 (1.54, 2.23) | 1.43 (1.36, 1.5) | 1.75 (1.46, 2.09) | 1.79 (1.5, 2.13) | 1.81 (1.51, 2.16) | 2.13 (1.8, 2.52) | 2.29 (1.94, 2.7) | 1.95 (1.62, 2.35) | 1.61 (1.52, 1.71) |
|  | Comorbidity Adjusted | 1.64 (1.39, 1.93) | 1.74 (1.48, 2.04) | 2.15 (1.86, 2.49) | 1.64 (1.38, 1.94) | 1.5 (1.25, 1.8) | 1.49 (1.23, 1.79) | 1.39 (1.33, 1.46) | 1.45 (1.21, 1.73) | 1.49 (1.25, 1.78) | 1.52 (1.27, 1.82) | 1.81 (1.52, 2.14) | 1.96 (1.66, 2.31) | 1.68 (1.4, 2.02) | 1.53 (1.45, 1.62) |
| Child's Childhood Onset Disorder |  |  |  |  |  |  |  |  |  |  |  |  |  |  |  |
|  | Minimally Adjusted | 2.1 (1.99, 2.23) | 2.56 (2.43, 2.7) | 2.57 (2.44, 2.71) | 1.98 (1.87, 2.1) | 1.95 (1.84, 2.07) | 1.76 (1.65, 1.88) | 1.52 (1.49, 1.55) | 2.29 (2.05, 2.56) | 2.21 (1.97, 2.48) | 2.66 (2.39, 2.96) | 2.33 (2.07, 2.62) | 2.25 (2, 2.54) | 2.01 (1.76, 2.29) | 1.75 (1.68, 1.83) |
|  | Comorbidity Adjusted | 1.96 (1.85, 2.07) | 2.4 (2.27, 2.52) | 2.42 (2.3, 2.55) | 1.88 (1.77, 1.99) | 1.86 (1.76, 1.98) | 1.69 (1.59, 1.8) | 1.57 (1.54, 1.61) | 2.05 (1.84, 2.3) | 1.99 (1.77, 2.23) | 2.41 (2.17, 2.68) | 2.13 (1.89, 2.39) | 2.07 (1.83, 2.33) | 1.85 (1.63, 2.11) | 1.73 (1.66, 1.8) |

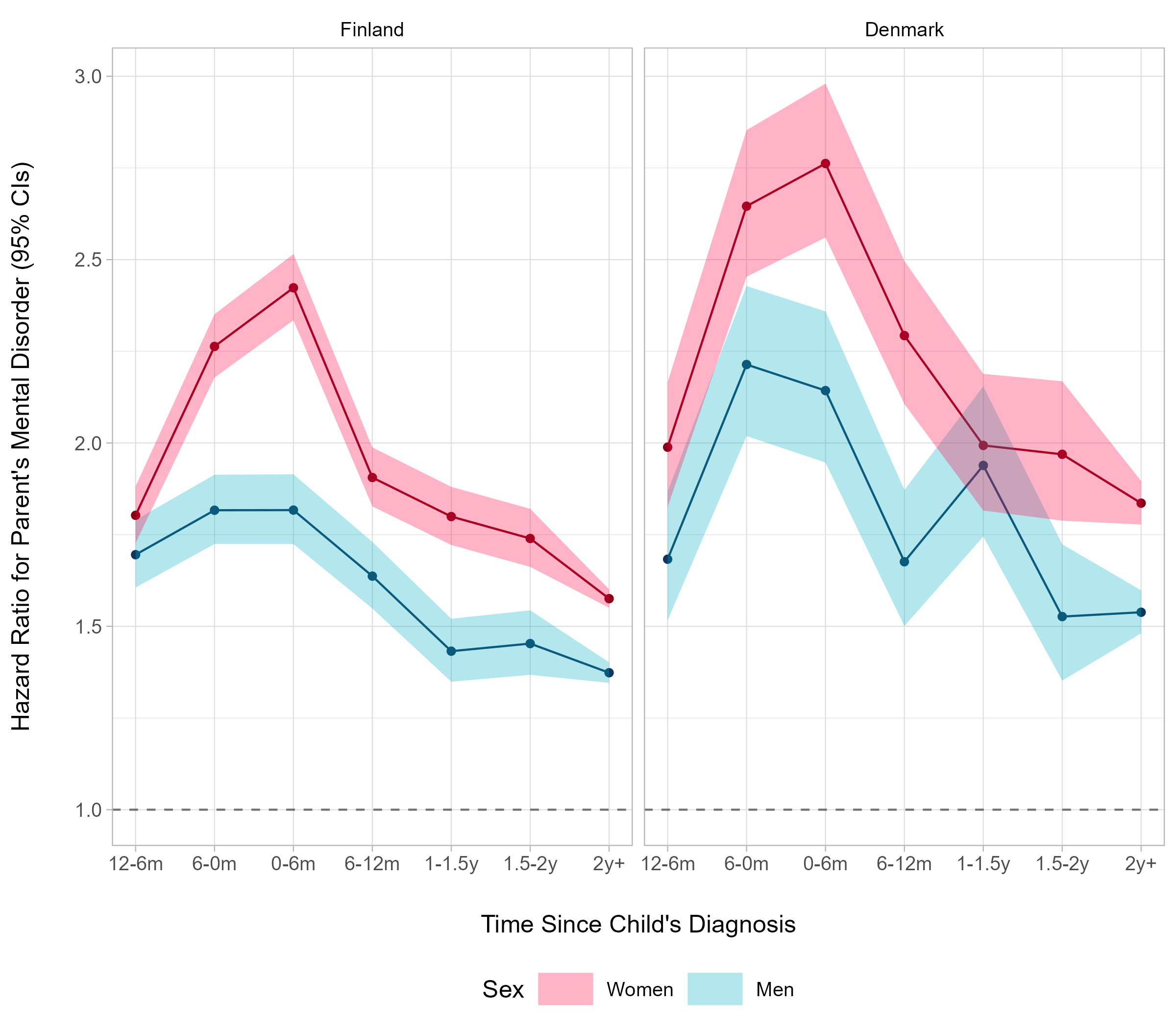

### **Supplement Figure 1.** Hazard Ratios and 95% Confidence Intervals of Parents’ Mental Disorder in Relation to Any Mental Disorder of a Child Before and After the Child’s Diagnosis

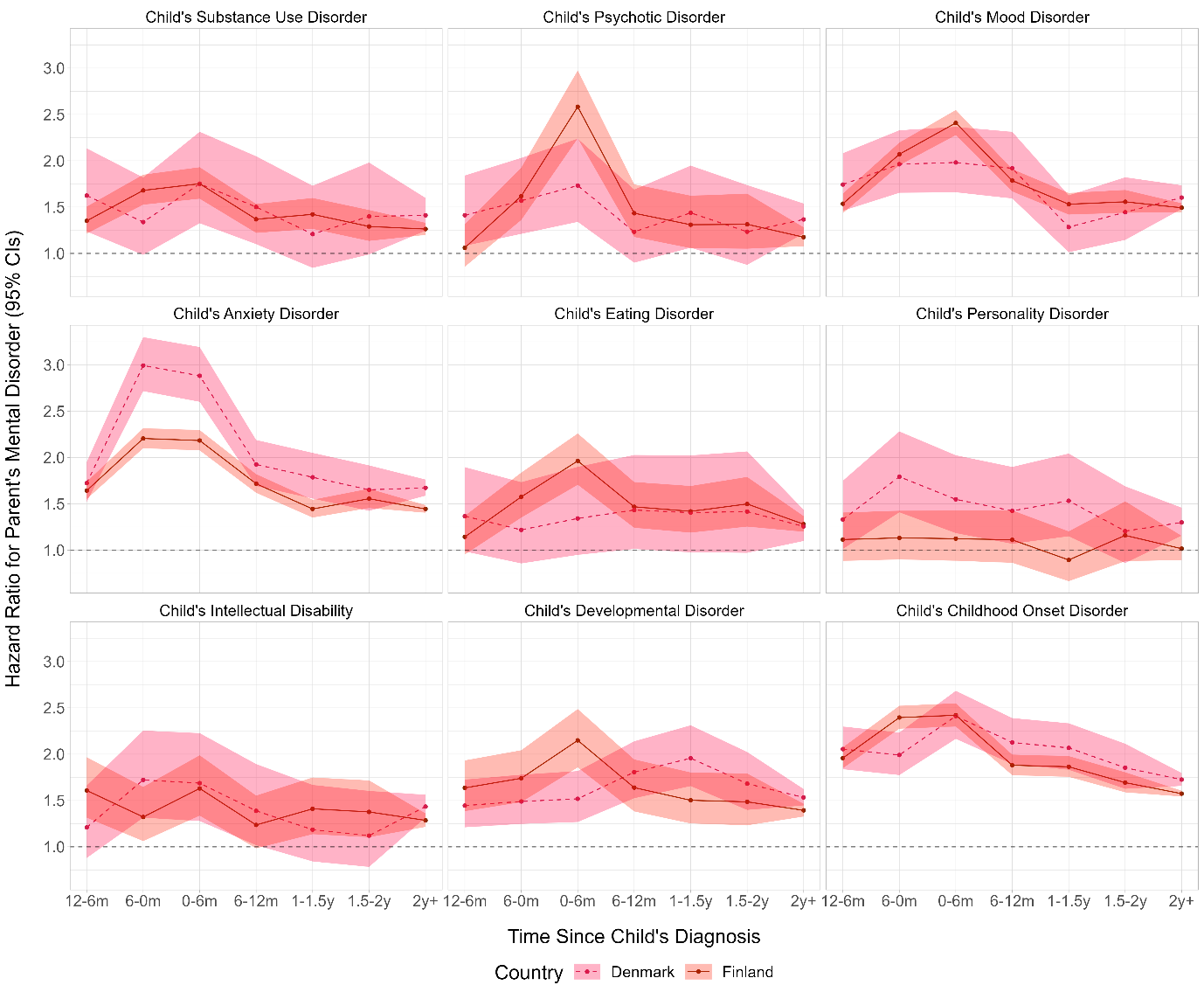

### **Supplement Figure 2.** Women's Hazard Ratios and 95% Confidence Intervals of Mental Disorder in Relation to a Child’s Specific Mental Disorder Before and After the Child’s Diagnosis

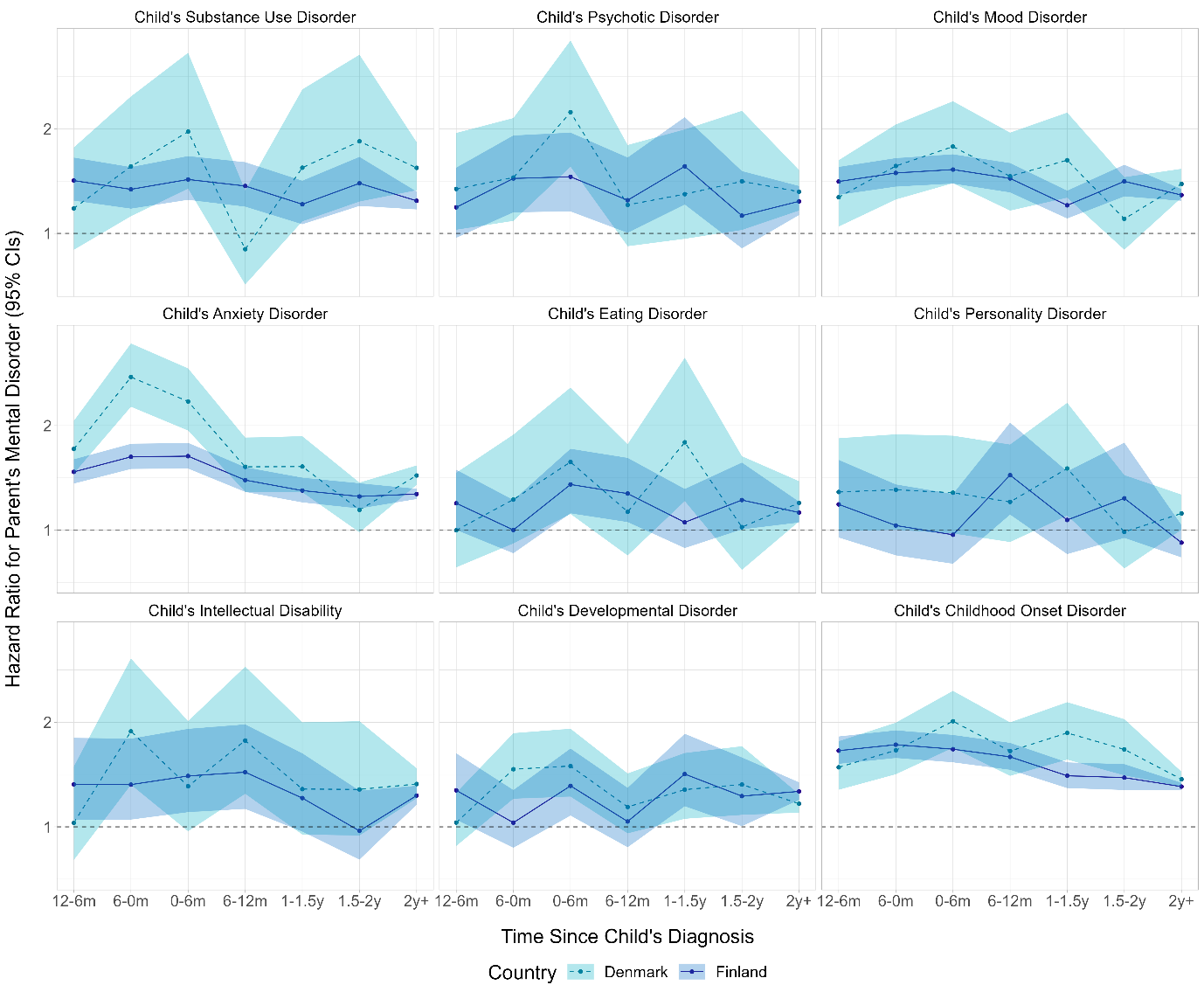

### **Supplement Figure 3.** Men's Hazard Ratios and 95% Confidence Intervals of Mental Disorder in Relation to a Child’s Specific Mental Disorder Before and After the Child’s Diagnosis

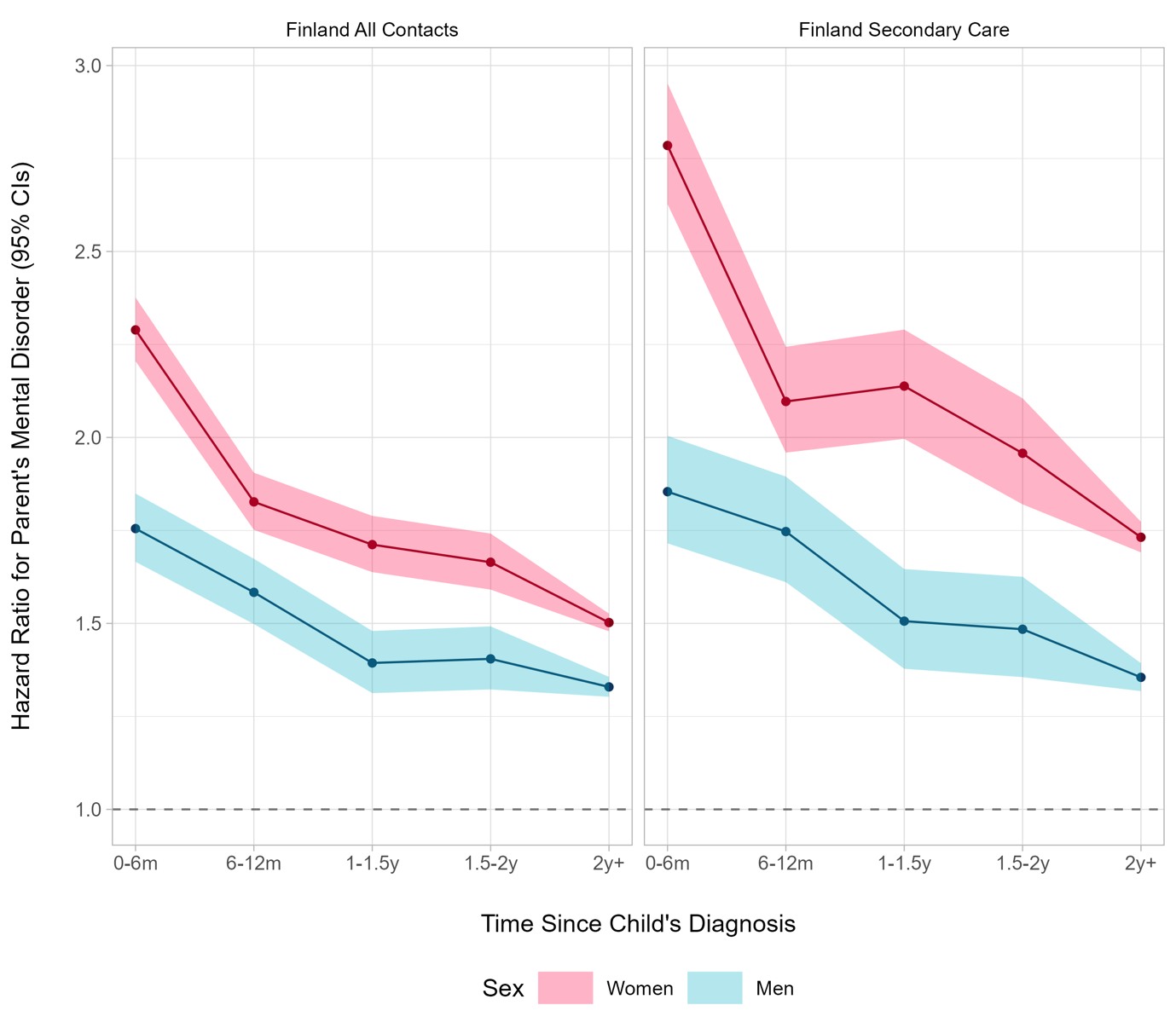

### **Supplement Figure 4.** Hazard Ratios and 95% Confidence Intervals of Parents’ Mental Disorder in Relation to Any Mental Disorder After the Child’s Diagnosis in Finland

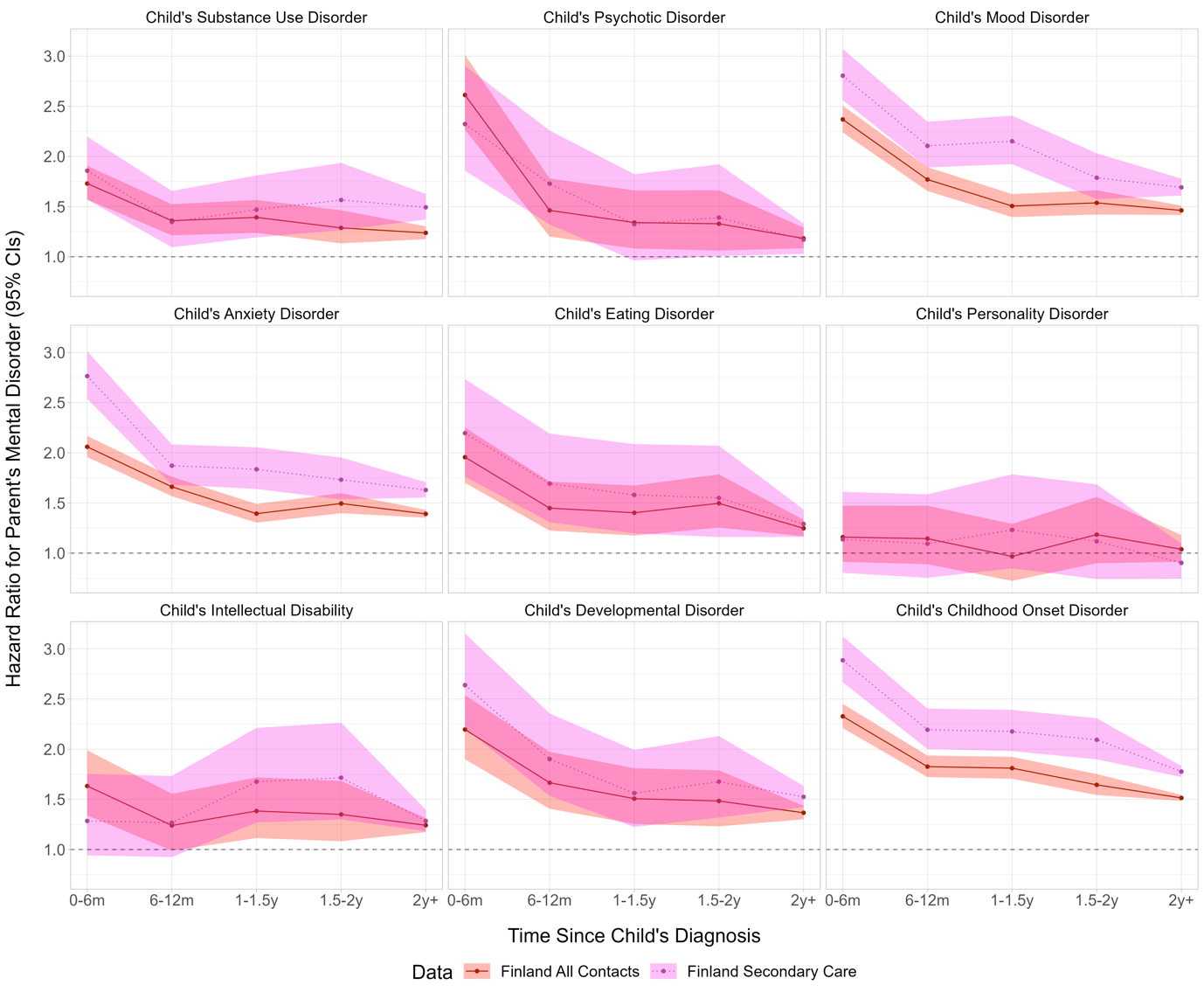

### **Supplement Figure 5.** Women's Hazard Ratios and 95% Confidence Intervals of Mental Disorder in Relation to a Child’s Specific Mental Disorder After the Child’s Diagnosis in Finland

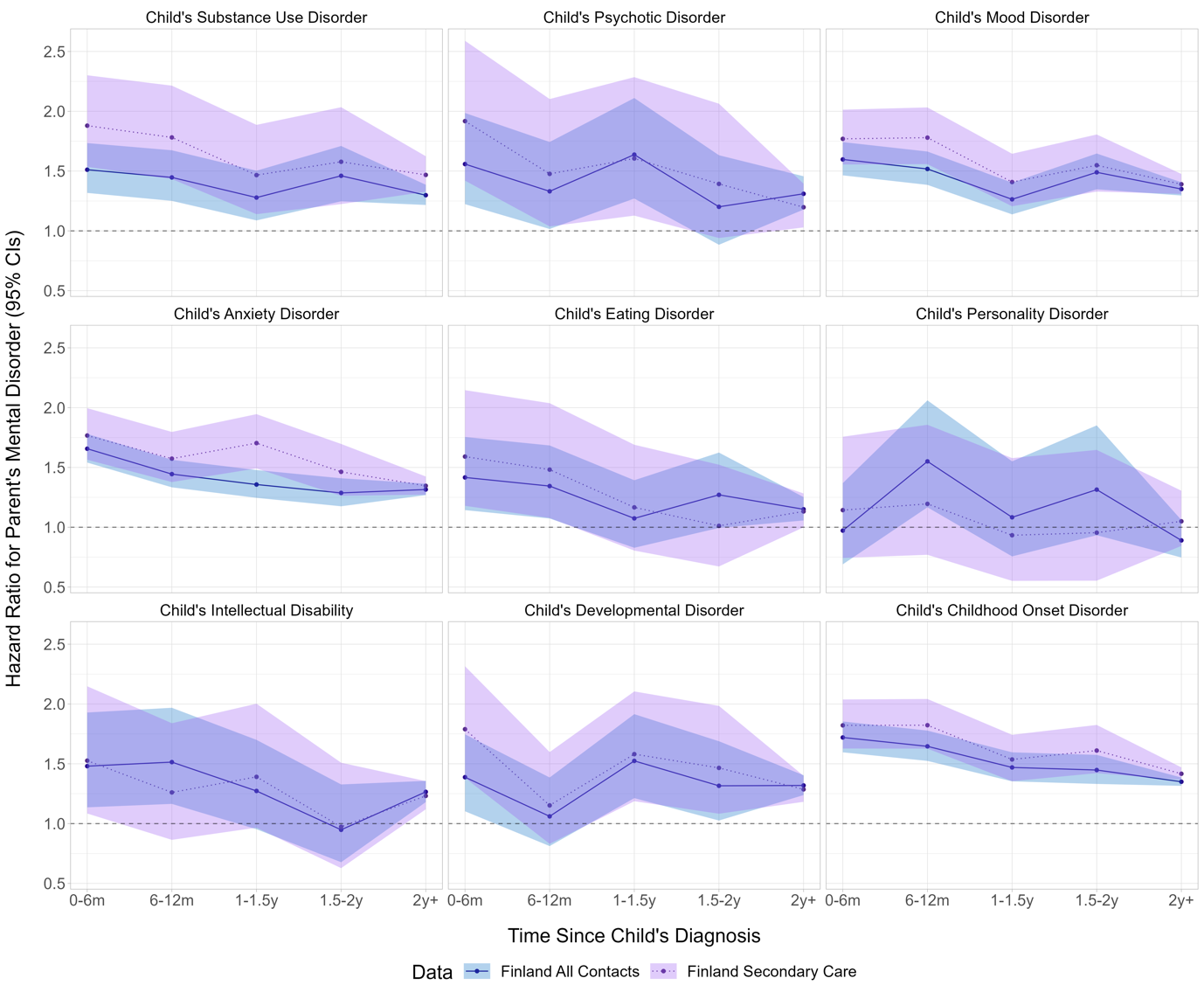

### **Supplement Figure 6.** Men's Hazard Ratios and 95% Confidence Intervals of Mental Disorder in Relation to a Child’s Specific Mental Disorder After the Child’s Diagnosis in Finland
